## Supplementary figures and tables for "Unravelling *Chlamydia trachomatis* Diversity in Amhara, Ethiopia: MLVA-*ompA* Sequencing as a Molecular Typing Tool for Trachoma"

**Supplementary tables and Figures**

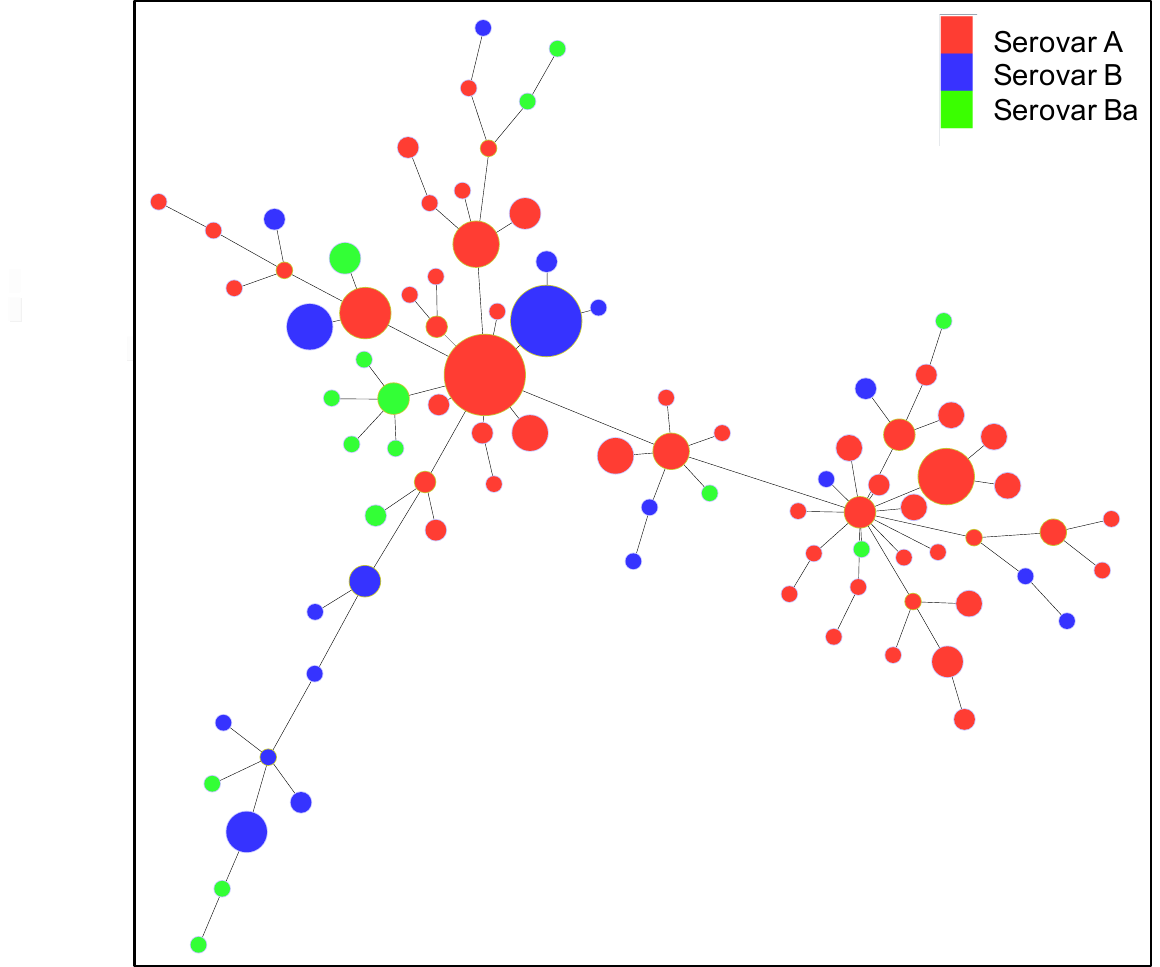

**Supplementary Fig. 1**. A minimum spanning tree showing the sequence-types identified in the Amhara region of Ethiopia between 2011-2017. Each MLVA-*omp*A sequence-type (n= 87) is coloured by serovar. Each circle represents a different sequence-type, with the size of circle being directly proportional to the number of individuals who had that variant. Trees were generated using Phyloviz 2.0.

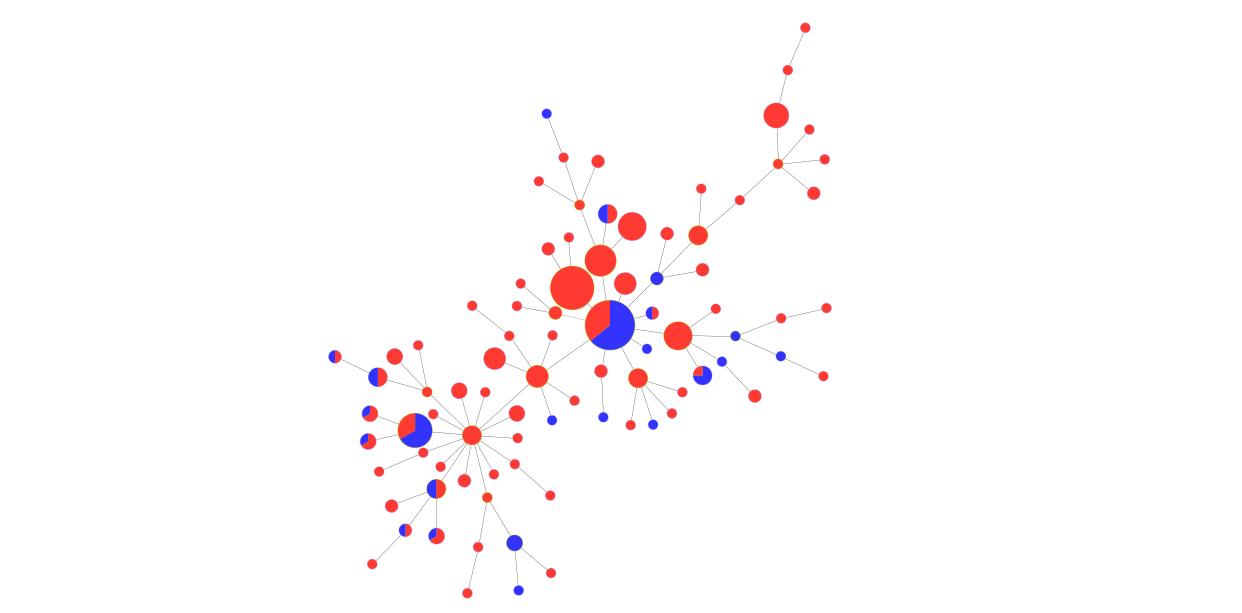

**Supplementary Fig. 2.** A minimum spanning tree showing the sequence-types identified in the Amhara region of Ethiopia between 2011-2017. Each MLVA-*omp*A sequence-type (n= 87) is coloured by number of rounds of MDA, either 5 rounds (red) or 8-10 rounds (blue). Each circle represents a different sequence-type, with the size of circle being directly proportional to the number of individuals who had that variant. Trees were generated using Phyloviz 2.0.

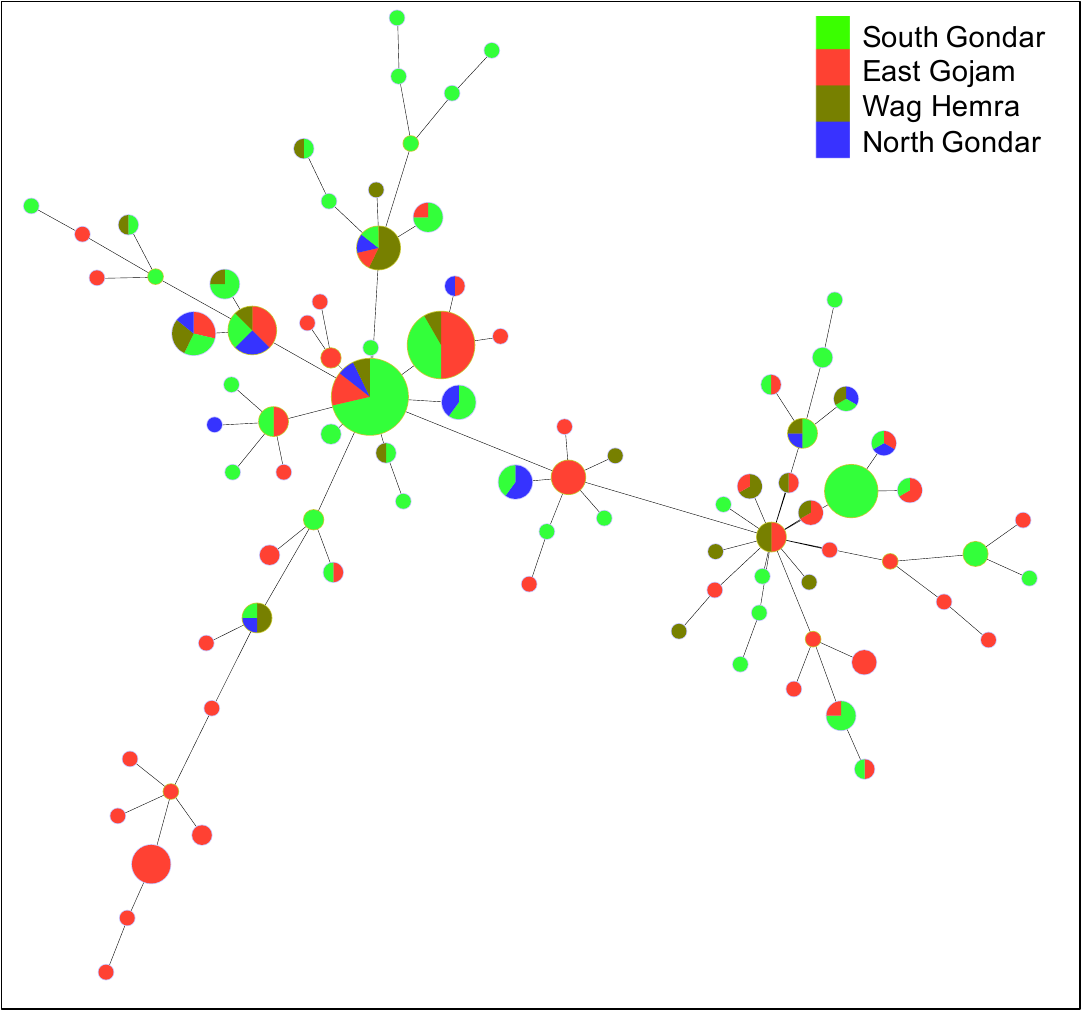

**Supplementary Fig. 3**. A minimum spanning tree showing the sequence-types identified in the Amhara region of Ethiopia between 2011-2017. Each MLVA-*omp*A sequence-type (n= 87) is coloured by zone. Each circle represents a different sequence-type, with the size of circle being directly proportional to the number of individuals who had that variant. Trees were generated using Phyloviz 2.0.

**Supplementary table 1**. The total number of Ethiopian conjunctival samples unable to be sequenced using Sanger sequencing for any of the three VNTRs and/or *omp*A for any reason (denoted as an “NA”), split by year of collection. VNTR: Variable number tandem repeats.

| **Year** | **Total samples** | **Total NAs** | **Proportion** |
| --- | --- | --- | --- |
| 2004 | 69 | 30 | 0.43 |
| 2005 | 51 | 23 | 0.45 |
| 2006 | 100 | 28 | 0.28 |
| 2007 | 56 | 18 | 0.32 |
| 2008 | 14 | 4 | 0.29 |
| 2010 | 10 | 3 | 0.30 |

**Supplementary table 2.** WGS results from 99 samples vs Sanger sequencing results for the three VNTRs CT1291, CT1299 and CT1335. WGS: Whole genome sequencing. VNTR: Variable number tandem repeat.

| **ID** | **CT1291_WGS** | **CT1291_Sanger** | **CT1335_WGS** | **CT1335_Sanger** | **CT1299_WGS** | **CT1299_Sanger** |
| --- | --- | --- | --- | --- | --- | --- |
| **1459-6700** | NA | AAATGGTCT_10C | GAAAAAGG_10T/8A | GAAAAGG_10T/8A | ATTCT_NA_ | ATTCT_10C_ |
| **1496-6885** | AAATGGTCT_9C | AAATGGTCT_NA | GAAAAAGG_10T/8A | GAAAAAGG_NA | ATTCT_14C_ | NA |
| **1524-7024** | AAATGGTCT_9C | AAATGGTCT_10C | GAAAAAGG_10T/8A | GAAAAGG_10T/8A | ATTCT_14C_ | ATTCT_10C_ |
| **1612-7464** | AAATGGTCT_10C | AAATGGTCT_9C | GAAAAAGG_10T/8A | GAAAAGG_10T/8A | NA | ATTCT_14C_ |
| **1623-7519** | AAATGGTCT_10C | AAATGGTCT_10C | GAAAAAGG_10T/8A | GAAAAAGG_10T/8A | NA | ATTCT_13C_ |
| **1629-7546** | AAATGGTCT_10C | AAATGGTCT_9C | GAAAAAGG_10T/8A | GAAAAGG_10T/8A | NA | ATTCT_10C_ |
| **1630-7550** | AAATGGTCT_9C | AAATGGTCT_9C | GAAAAAGG_10T/8A | GAAAAGG_10T/8A | ATTCT_14C_ | ATTCT_11C_ |
| **1633-7568** | AAATGGTCT_9C | AAATGGTCT_9C | GAAAAAGG_10T/8A | GAAAAGG_10T/8A | NA | ATTCT_11C_ |
| **1653-7666** | AAATGGTCT_10C | AAATGGTCT_NA | GAAAAAGG_10T/8A | GAAAAAGG_10T/8A | NA | ATTCT_11C |
| **1658-7694** | AAATGGTCT_9C | AAATGGTCT_8C | GAAAAAGG_10T/8A | GAAAAGG_10T/8A | NA | ATTCT_14C_ |
| **1667-7738** | AAATGGTCT_10C | AAATGGTCT_9C | GAAAAAGG_10T/8A | GAAAAGG_10T/8A | ATTCT_14C_ | ATTCT_10C_ |
| **1675-7776** | AAATGGTCT_9C | AAATGGTCT_9C | GAAAAAGG_10T/8A | GAAAAGG_10T/8A | ATTCT_14C_ | ATTCT_12C_ |
| **1680-7799** | AAATGGTCT_9C | AAATGGTCT_9C | GAAAAAGG_10T/8A | GAAAAGG_10T/8A | ATTCT_14C_ | ATTCT_10C_ |
| **1682-7810** | AAATGGTCT_10C | AAATGGTCT_9C | GAAAAAGG_10T/8A | GAAAAAGG_10T/8A | NA | ATTCT_11C_ |
| **1689-7847** | AAATGGTCT_10C | AAATGGTCT_9C | GAAAAAGG_10T/8A | GAAAAAGG_10T/8A | ATTCT_14C_ | ATTCT_10C_ |
| **1710-7953** | AAATGGTCT_9C | AAATGGTCT_9C | GAAAAAGG_10T/8A | GAAAAAGG_NA | NA | ATTCT_10C |
| **1767-8232** | AAATGGTCT_9C | AAATGGTCT_10C | GAAAAAGG_10T/8A | GAAAAGG_10T/8A | NA | ATTCT_14C_ |
| **1770-8246** | NA | AAATGGTCT_NA | GAAAAAGG_10T/8A | GAAAAAGG_10T/8A | NA | ATTCT_14C |
| **1773-8261** | AAATGGTCT_9C | AAATGGTCT_9C | GAAAAAGG_10T/8A | GAAAAGG_10T/8A | ATTCT_14C_ | ATTCT_11C_ |
| **1781-8301** | AAATGGTCT_9C | AAATGGTCT_9C | GAAAAAGG_10T/8A | GAAAAAGG_10T/8A | ATTCT_14C_ | ATTCT_15C |
| **1801-8400** | AAATGGTCT_9C | AAATGGTCT_10C | GAAAAAGG_10T/8A | GAAAAGG_10T/8A | ATTCT_14C_ | ATTCT_11C_ |
| **1859-8691** | AAATGGTCT_10C | AAATGGTCT_9C | GAAAAAGG_10T/8A | GAAAAGG_10T/8A | ATTCT_14C_ | ATTCT_10C_ |
| **1866-8726** | AAATGGTCT_9C | AAATGGTCT_8C | GAAAAAGG_10T/8A | GAAAAAGG_10T/8A | ATTCT_14C_ | NA |
| **1870-8742** | AAATGGTCT_9C | AAATGGTCT_9C | GAAAAAGG_10T/8A | GAAAAAGG_10T/8A | NA | ATTCT_15C |
| **1878-8783** | AAATGGTCT_NA | AAATGGTCT_9C | GAAAAAGG_10T/8A | GAAAAGG_10T/8A | NA | ATTCT_10C_ |
| **1879-8790** | AAATGGTCT_10C | AAATGGTCT_9C | GAAAAAGG_10T/8A | GAAAAGG_10T/8A | ATTCT_14C_ | ATTCT_10C_ |
| **2134-9948** | AAATGGT_12C | AAATGGT_8C | GAAAAAGG_10T/8A | GAAAAGG_10T/8A | ATTCTATTCT_9C_ | ATTCT_9C_ |
| **2136-9960** | AAATGGTCT_9C | AAATGGTCT_9C | GAAAAAGG_10T/8A | GAAAAGG_10T/8A | NA | ATTCT_11C_ |
| **2137-9962** | NA | AAATGGT_8C | GAAAAAGG_10T/8A | GAAAAAGG_10T/8A | ATTCT_14C_ | ATTCT_9C |
| **2148-10019** | AAATGGTCT_9C | AAATGGTCT_9C | GAAAAAGG_10T/8A | GAAAAGG_10T/8A | ATTCT_14C_ | ATTCT_10C_ |
| **2149-10022** | AAATGGTCT_9C | AAATGGTCT_8C | GAAAAAGG_10T/8A | GAAAAGG_10T/8A | ATTCT_14C_ | ATTCT_10C_ |
| **2166-10103** | AAATGGTCT_9C | AAATGGTCT_8C | GAAAAAGG_10T/8A | GAAAAGG_10T/8A | ATTCT_14C_ | ATTCT_16C |
| **2166-10106** | AAATGGTCT_9C | AAATGGTCT_9C | GAAAAAGG_10T/8A | GAAAAGG_10T/8A | ATTCT_14C_ | ATTCT_11C_ |
| **2169-10121** | AAATGGTCT_9C | AAATGGTCT_8C | GAAAAAGG_10T/8A | GAAAAGG_10T/8A | ATTCT_14C_ | ATTCT_10C_ |
| **2174-10147** | AAATGGTCT_9C | AAATGGTCT_8C | GAAAAAGG_10T/8A | GAAAAAGG_10T/8A | NA | ATTCT_14C |
| **2175-10152** | AAATGGTCT_9C | AAATGGTCT_8C | GAAAAAGG_10T/8A | GAAAAGG_10T/8A | ATTCT_3CT_10C_ | ATTCT_10C_ |
| **2178-10163** | AAATGGTCT_9C | AAATGGTCT_9C | GAAAAAGG_NT/8A | GAAAAGG_9T/8A | ATTCT_14C_ | ATTCT_9C_ |
| **2182-10186** | AAATGGTCT_9C | AAATGGTCT_8C | GAAAAAGG_10T/8A | GAAAAGG_10T/8A | ATTCT_14C_ | ATTCT_14C_ |
| **2188-10216** | AAATGGTCT_NA | AAATGGTCT_8C | GAAAAAGG_10T/8A | GAAAAGG_10T/8A | ATTCT_14C_ | ATTCT_13C_ |
| **2209-10263** | AAATGGTCT_NA | AAATGGTCT_8C | GAAAAAGG_NT/8A | GAAAAGG_9T/8A | ATTCT_14C_ | ATTCT_11C_ |
| **2228-10360** | AAATGGTCT_NA | AAATGGTCT_8C | GAAAAAGG_10T/8A | GAAAAGG_10T/8A | ATTCT_14C_ | ATTCT_10C_ |
| **2233-10384** | AAATGGTCT_NA | AAATGGTCT_8C | GAAAAAGG_10T/8A | GAAAAGG_10T/8A | ATTCT_14C_ | ATTCT_10C_ |
| **2241-10425** | AAATGGT_12C | AAATGGT_8C | GAAAAAGG_10T/8A | GAAAAAGG_NA | NA | ATTCT_9C |
| **2243-10433** | AAATGGT_NA | AAATGGTCT_8C | GAAAAAGG_10T/8A | GAAAAAGG_10T/8A | ATTCTCTTCT_9C_ | ATTCT_10C_ |
| **2243-10434** | AAATGGT_12C | AAATGGT_8C | GAAAAAGG_10T/8A | GAAAAGG_10T/8A | ATTCTATTCT_9C_ | ATTCT_9C_ |
| **2247-10454** | AAATGGT_NA | AAATGGT_8C | GAAAAAGG_10T/8A | GAAAAGG_10T/8A | ATTCT_14C_ | ATTCT_10C_ |
| **2247-10456** | AAATGGTCT_10C | AAATGGTCT_9C | GAAAAAGG_10T/8A | GAAAAGG_10T/8A | ATTCT_14C_ | ATTCT_10C_ |
| **2248-10461** | NA | AAATGGTCT_9C | GAAAAAGG_10T/8A | GAAAAGG_10T/8A | NA | ATTCT_10C_ |
| **2251-10472** | AAATGGT_NA | AAATGGT_9C | GAAAAAGG_10T/8A | GAAAAAGG_10T/8A | NA | ATTCT_10C_ |
| **2254-10487** | AAATGGT_12C | AAATGGT_8C | GAAAAAGG_10T/8A | GAAAAGG_10T/8A | ATTCT_14C_ | ATTCT_9C_ |
| **2255-10495** | AAATGGT_NA | AAATGGT_8C | GAAAAAGG_10T/8A | GAAAAGG_10T/8A | ATTCTATTCT_9C_ | ATTCT_9C_ |
| **2256-10501** | AAATGGT_12C | AAATGGT_8C | GAAAAAGG_10T/8A | GAAAAGG_10T/8A | NA | ATTCT_9C_ |
| **2258-10504** | AAATGGTCT_9C | AAATGGTCT_9C | GAAAAAGG_10T/8A | GAAAAGG_10T/8A | NA | ATTCT_10C_ |
| **2268-10556** | AAATGGTCT_9C | AAATGGTCT_9C | GAAAAAGG_NT/8A | GAAAAGG_9T/8A | NA | ATTCT_9C_ |
| **2280-10615** | AAATGGTCT_10C | AAATGGTCT_9C | GAAAAAGG_10T/8A | GAAAAGG_10T/8A | NA | ATTCT_11C_ |
| **2294-10681** | AAATGGTCT_9C | AAATGGTCT_7C | GAAAAAGG_10T/8A | GAAAAAGG_10T/8A | ATTCT_14C_ | ATTCT_11C_ |
| **2303-10728** | AAATGGTCT_9C | AAATGGTCT_8C | GAAAAAGG_10T/8A | GAAAAGG_10T/8A | ATTCT_14C_ | ATTCT_12C_ |
| **2314-10783** | AAATGGTCT_NA | AAATGGTCT_9C | GAAAAAGG_10T/8A | GAAAAGG_10T/8A | ATTCT_14C_ | ATTCT_10C_ |
| **2356-10989** | AAATGGTCT_9C | AAATGGTCT_9C | GAAAAAGG_10T/8A | GAAAAGG_10T/8A | ATTCT_14C_ | ATTCT_10C_ |
| **2394-11155** | AAATGGTCT_9C | AAATGGTCT_9C | GAAAAAGG_10T/8A | GAAAAGG_10T/8A | NA | ATTCT_10C_ |
| **2405-11212** | NA | AAATGGTCT_11C | GAAAAAGG_10T/8A | GAAAAGG_10T/8A | NA | ATTCT_14C_ |
| **2406-11215** | NA | AAATGGTCT_NA | GAAAAAGG_10T/8A | GAAAAGG_10T/8A | NA | ATTCT_14C |
| **2437-11366** | NA | AAATGGTCT_11C | GAAAAAGG_10T/8A | GAAAAGG_10T/8A | ATTCT_14C_ | ATTCT_14C_ |
| **2441-11387** | AAATGGTCT_9C | AAATGGTCT_9C | GAAAAAGG_10T/8A | GAAAAGG_10T/8A | NA | ATTCT_11C_ |
| **2451-11436** | AAATGGTCT_10C | AAATGGTCT_9C | GAAAAAGG_10T/8A | GAAAAGG_10T/8A | ATTCT_14C_ | ATTCT_10C_ |
| **2452-11442** | AAATGGTCT_10C | AAATGGTCT_9C | GAAAAAGG_10T/8A | GAAAAAGG_10T/8A | ATTCT_14C_ | ATTCT_9C_ |
| **2495-11650** | AAATGGTCT_9C | AAATGGT_9C | GAAAAAGG_10T/8A | GAAAAGG_10T/8A | ATTCT_14C_ | ATTCT_10C_ |
| **2495-11652** | AAATGGT_12C | AAATGGTCT_8C | GAAAAAGG_10T/8A | GAAAAAGG_NA | ATTCT_14C_ | ATTCT_9C |
| **2496-11655** | AAATGGTCT_10C | AAATGGTCT_9C | GAAAAAGG_10T/8A | GAAAAAGG_NA | ATTCT_14C_ | ATTCT_10C |
| **2972-13924** | AAATGGTCT_10C | AAATGGTCT_NA | GAAAAAGG_10T/8A | GAAAAAGG_NA | ATTCT_14C_ | NA |
| **2976-13947** | AAATGGTCT_9C | AAATGGTCT_9C | GAAAAAGG_10T/8A | GAAAAGG_10T/8A | ATTCT_14C_ | ATTCT_11C_ |
| **2983-13981** | AAATGGTCT_10C | AAATGGTCT_9C | GAAAAAGG_10T/8A | GAAAAAGG_10T/8A | ATTCT_14C_ | ATTCT_10C_ |
| **2987-13999** | AAATGGTCT_9C | AAATGGTCT_9C | GAAAAAGG_10T/8A | GAAAAGG_10T/8A | ATTCT_14C_ | ATTCT_11C_ |
| **2991-14016** | AAATGGTCT_10C | AAATGGTCT_9C | GAAAAAGG_10T/8A | GAAAAGG_10T/8A | ATTCT_14C_ | ATTCT_11C_ |
| **2994-14032** | AAATGGTCT_9C | AAATGGTCT_9C | GAAAAAGG_10T/8A | GAAAAGG_10T/8A | ATTCT_14C_ | ATTCT_10C_ |
| **3002-14069** | NA | AAATGGTCT_10C | GAAAAAGG_10T/8A | GAAAAGG_10T/8A | ATTCT_14C_ | ATTCT_10C_ |
| **3004-14083** | NA | AAATGGT_8C | GAAAAAGG_10T/8A | GAAAAGG_10T/8A | ATTCTCTTCT_9C_ | ATTCT_10C_ |
| **3009-14104** | AAATGGTCT_9C | AAATGGTCT_9C | GAAAAAGG_10T/8A | GAAAAAGG_10T/8A | NA | ATTCT_15C |
| **3027-14195** | AAATGGTCT_9C | AAATGGTCT_9C | GAAAAAGG_10T/8A | GAAAAGG_10T/8A | NA | ATTCT_13C_ |
| **3030-14213** | AAATGGTCT_9C | AAATGGTCT_9C | GAAAAAGG_10T/8A | GAAAAGG_10T/8A | NA | ATTCT_15C |
| **3037-14244** | AAATGGTCT_9C | AAATGGTCT_9C | GAAAAAGG_10T/8A | GAAAAAGG_NA | NA | ATTCT_14C |
| **32-154** | AAATGGTCT_10C | AAATGGTCT_9C | GAAAAAGG_10T/8A | GAAAAAGG_10T/8A | ATTCT_14C_ | ATTCT_10C_ |
| **42-207** | AAATGGTCT_10C | AAATGGTCT_9C | GAAAAAGG_10T/8A | GAAAAGG_10T/8A | ATTCT_14C_ | ATTCT_10C_ |
| **45-216** | AAATGGTCT_9C | AAATGGTCT_9C | GAAAAAGG_10T/8A | GAAAAGG_10T/8A | ATTCT_14C_ | ATTCT_13C_ |
| **45-218** | AAATGGTCT_9C | AAATGGTCT_9C | GAAAAAGG_10T/8A | GAAAAGG_10T/8A | ATTCT_14C_ | ATTCT_11C_ |
| **62-303** | AAATGGTCT_9C | AAATGGTCT_8C | GAAAAAGG_10T/8A | GAAAAAGG_10T/8A | NA | ATTCT_18C |
| **64-311** | AAATGGTCT_9C | AAATGGTCT_8C | GAAAAAGG_10T/8A | GAAAAGG_10T/8A | NA | ATTCT_14C_ |
| **65-315** | AAATGGTCT_9C | AAATGGTCT_8C | GAAAAAGG_10T/8A | GAAAAAGG_10T/8A | NA | ATTCT_16C |
| **71-349** | AAATGGTCT_9C | AAATGGTCT_12C | GAAAAAGG_10T/8A | GAAAAAGG_NA | ATTCT_14C_ | ATTCT_14C |
| **75-367** | AAATGGTCT_9C | AAATGGTCT_12C | GAAAAAGG_10T/8A | GAAAAGG_10T/8A | ATTCT_14C_ | ATTCT_10C_ |
| **850-3968** | AAATGGTCT_9C | AAATGGTCT_9C | GAAAAAGG_10T/8A | GAAAAGG_10T/8A | ATTCT_14C_ | ATTCT_10C_ |
| **865-4011** | AAATGGTCT_9C | AAATGGTCT_9C | GAAAAAGG_10T/8A | GAAAAGG_10T/8A | ATTCT_14C_ | ATTCT_11C_ |
| **865-4012** | AAATGGTCT_10C | AAATGGTCT_9C | GAAAAAGG_10T/8A | GAAAAAGG_NA | NA | ATTCT_11C |
| **866-4014** | AAATGGTCT_10C | AAATGGTCT_9C | GAAAAAGG_10T/8A | GAAAAAGG_10T/8A | ATTCT_14C_ | ATTCT_10C_ |
| **872-4044** | AAATGGTCT_10C | AAATGGTCT_9C | GAAAAAGG_10T/8A | GAAAAGG_10T/8A | NA | ATTCT_11C_ |
| **873-4051** | AAATGGTCT_10C | AAATGGTCT_9C | GAAAAAGG_10T/8A | GAAAAAGG_NA | ATTCT_14C_ | ATTCT_11C |
| **884-4103** | AAATGGTCT_10C | AAATGGTCT_9C | GAAAAAGG_10T/8A | GAAAAAGG_10T/8A | ATTCT_14C_ | ATTCT_11C_ |
| **889-4129** | AAATGGTCT_9C | AAATGGTCT_9C | GAAAAAGG_10T/8A | GAAAAGG_10T/8A | ATTCT_14C_ | ATTCT_13C_ |
| **889-4130** | AAATGGTCT_9C | AAATGGTCT_9C | GAAAAAGG_10T/8A | GAAAAGG_10T/8A | ATTCT_14C_ | ATTCT_13C_ |

**Supplementary table 3**. The serovar and Pedersen sequence-type associated with each sample ID used to generate the phylogenetic trees (n=45).

| **ID** | **Pedersen Code** | **Serovar** |
| --- | --- | --- |
| 1524-7024 | 3b.4.4 | A |
| 1675-7776 | 3b.6.3 | A |
| 1680-7799 | 3b.4.3 | A |
| 1773-8261 | 3b.5.3 | A |
| 1801-8400 | 3b.5.4 | A |
| 2148-10019 | 3b.4.3 | A |
| 2149-10022 | 3b.4.2 | A |
| 2166-10106 | 3b.5.3 | A |
| 2169-10121 | 3b.4.2 | A |
| 2175-10152 | 3b.4.2 | A |
| 2178-10163 | 1a.3.3 | A |
| 2182-10186 | 3b.8.2 | A |
| 2294-10681 | 3.5.8 | A |
| 2303-10728 | 3b.6.2 | A |
| 2976-13947 | 3b.5.3 | A |
| 2987-13999 | 3b.5.3 | A |
| 2994-14032 | 3b.4.3 | A |
| 45-216 | 3b.7.3 | A |
| 45-218 | 3b.5.3 | A |
| 71-347 | 3.8.2 | A |
| 75-367 | 3b.4.7 | A |
| 850-3968 | 3b.4.3 | A |
| 889-4129 | 3b.7.3 | A |
| 889-4130 | 3b.7.3 | A |
| 1630-7550 | 3b.5.3 | B |
| 1667-7738 | 3b.4.3 | B |
| 1689-7847 | 3.4.3 | B |
| 1859-8691 | 3b.4.3 | B |
| 1879-8790 | 3b.4.3 | B |
| 2134-9948 | 3b.3.2c | B |
| 2243-10434 | 3b.3.2c | B |
| 2254-10487 | 3b.3.2c | B |
| 2356-10989 | 3b.4.3 | B |
| 2451-11436 | 3b.4.3 | B |
| 2452-11442 | 3.3.3 | B |
| 2495-11650 | 3b.4.3c | B |
| 2983-13981 | 3.4.3 | B |
| 2991-14016 | 3b.5.3 | B |
| 32-154 | 3.4.3 | B |
| 42-207 | 3b.4.3 | B |
| 865-4011 | 3b.5.3 | B |
| 866-4014 | 3.4.3 | B |
| 884-4103 | 3.5.3 | B |
