## Supplementary material for "Unravelling *Chlamydia trachomatis* Diversity in Amhara, Ethiopia: MLVA-*ompA* Sequencing as a Molecular Typing Tool for Trachoma": Data

| ID | Age | sex_sv | elembod_count | survey_ind | TFgott | Tigott |
|---|---|---|---|---|---|---|
| 1459-6700 | 1-3 | 2. Female | 219.8313904 | 2 | 40.625 | 18.76 |
| 1524-7024 | 4-5 | 2. Female | 156.2561951 | 2 | 33.3333333 | 22.2322222 |
| 1541-7108 | 4-5 | 2. Female | 37.98825073 | 2 | 36.8421053 | 3.51877193 |
| 1548-7144 | 4-5 | 1. Male | 25.71686745 | 2 | 13.4615385 | 1.93307692 |
| 1557-7190 | 4-5 | 1. Male | 9.602944374 | 2 | 36.8421053 | 3.51877193 |
| 1560-7203 | 1-3 | 1. Male | 14.74954987 | 2 | 33.3333333 | 22.2322222 |
| 1612-7464 | 4-5 | 1. Male | 29.19330406 | 2 | 41.4634146 | 14.6441463 |
| 1614-7470 | 4-5 | 2. Female | 30.3547287 | 2 | 61.4285714 | 10.01 |
| 1620-7502 | 4-5 | 1. Male | 2914.547363 | 2 | 43.2432432 | 24.3343243 |
| 1621-7508 | 1-3 | 2. Female | 61.86403656 | 2 | 43.2432432 | 24.3343243 |
| 1623-7515 | 4-5 | 1. Male | 122.4454803 | 2 | 43.2432432 | 24.3343243 |
| 1623-7519 | 1-3 | 1. Male | 2517.877441 | 2 | 43.2432432 | 24.3343243 |
| 1629-7546 | 4-5 | 1. Male | 85.35303497 | 2 | 15.2542373 | 11.8744068 |
| 1630-7550 | 4-5 | 1. Male | 781.1620483 | 2 | 30.7692308 | 2.57410256 |
| 1633-7568 | 1-3 | 1. Male | 1738.100586 | 2 | 30.7692308 | 2.57410256 |
| 1641-7605 | 4-5 | 2. Female | 14.74954987 | 2 | 61.4285714 | 10.01 |
| 1647-7637 | 4-5 | 1. Male | 5.616121292 | 2 | 43.2432432 | 24.3343243 |
| 1655-7675 | 1-3 | 2. Female | 3.728496075 | 2 | 41.6666667 | 2.09333333 |
| 1658-7694 | 4-5 | 1. Male | 288.8626709 | 2 | 25.4237288 | 18.6540678 |
| 1667-7738 | 4-5 | 2. Female | 58.91953278 | 2 | 66.6666667 | 0.01 |
| 1675-7776 | 4-5 | 1. Male | 78.9464798 | 2 | 59.2592593 | 14.8248148 |
| 1680-7799 | 1-3 | 2. Female | 23.55570984 | 2 | 44.6428571 | 33.9385714 |
| 1682-7810 | 4-5 | 2. Female | 131.097229 | 2 | 44.6428571 | 33.9385714 |
| 1689-7847 | 1-3 | 1. Male | 781.1620483 | 2 | 83.3333333 | 0.01 |
| 1701-7907 | 1-3 | NA | 5.726747513 | 2 | 83.3333333 | 0.01 |
| 1708-7939 | 4-5 | 2. Female | 3.516568184 | 2 | 59.2592593 | 14.8248148 |
| 1767-8232 | 4-5 | 1. Male | 29.47941971 | 2 | 58.6956522 | 36.9665217 |
| 1773-8261 | 4-5 | 2. Female | 167.296936 | 2 | 61.8181818 | 5.46454546 |
| 1797-8381 | 1-3 | 2. Female | 154359.625 | 2 | 39.2857143 | 17.8671429 |
| 1801-8400 | 4-5 | 2. Female | 357.9963989 | 2 | 58.6956522 | 36.9665217 |
| 1805-8424 | 1-3 | 2. Female | 312.3040466 | 2 | 58.6956522 | 36.9665217 |
| 1857-8680 | 4-5 | 2. Female | 33.79247284 | 2 | 76.9230769 | 35.9074359 |
| 1859-8691 | 4-5 | 1. Male | 61.26358414 | 2 | 76.9230769 | 35.9074359 |
| 1878-8783 | 4-5 | 1. Male | 48.00737 | 2 | 37.7777778 | 17.7877778 |
| 1879-8790 | 4-5 | 2. Female | 36.89281845 | 2 | 76.9230769 | 35.9074359 |
| 1889-8840 | 4-5 | 2. Female | 17.07321358 | 2 | 29.4117647 | 11.7747059 |
| 1890-8842 | 4-5 | 1. Male | 7.5250597 | 2 | 29.4117647 | 11.7747059 |
| 1897-8878 | 4-5 | 1. Male | 4.804675579 | 2 | 76.9230769 | 35.9074359 |
| 1899-8888 | 1-3 | 1. Male | 560.6922607 | 2 | 76.9230769 | 35.9074359 |
| 2134-9948 | 1-3 | 2. Female | 1199.817627 | 5 | 60.7843137 | 5.89235294 |
| 2135-9954 | 4-5 | 2. Female | 913.0899048 | 5 | 60.7843137 | 5.89235294 |
| 2136-9960 | 4-5 | 2. Female | 688.1386108 | 5 | 67.6470588 | 41.1864706 |
| 2137-9963 | 1-3 | 2. Female | 27.00206947 | 5 | 67.6470588 | 41.1864706 |
| 2140-9981 | 1-3 | 1. Male | 10.0828476 | 5 | 67.6470588 | 41.1864706 |
| 2148-10018 | 1-3 | 2. Female | 10.18167114 | 5 | 43.8596491 | 1.76438597 |

| | | | | | | |
|---|---|---|---|---|---|---|
| 2148-10019 | 1-3 | 2. Female | 249.5484467 | 5 | 32.2580645 | 16.1390323 |
| 2149-10022 | 1-3 | 2. Female | 28.62936401 | 5 | 32.2580645 | 16.1390323 |
| 2150-10027 | 4-5 | 2. Female | 7.5250597 | 5 | 46.6666667 | 20.01 |
| 2166-10106 | 4-5 | 1. Male | 2775.825439 | 5 | 38.4615385 | 0.01 |
| 2168-10116 | 4-5 | 2. Female | 3.656471014 | 5 | 62.5 | 2.51 |
| 2169-10121 | 1-3 | 1. Male | 2196.511719 | 5 | 62.5 | 2.51 |
| 2174-10146 | 4-5 | 1. Male | 14.0475235 | 5 | 62.5 | 2.51 |
| 2175-10149 | 1-3 | 1. Male | 5.507632256 | 5 | 62.5 | 2.51 |
| 2175-10151 | 1-3 | 1. Male | 4.444038868 | 5 | 17.2413793 | 0.01 |
| 2175-10152 | 1-3 | 1. Male | 18593.85742 | 5 | 62.5 | 2.51 |
| 2178-10163 | 4-5 | 1. Male | 61.26358414 | 5 | 61.7021277 | 8.5206383 |
| 2182-10186 | 1-3 | 2. Female | 78.9464798 | 5 | 38.4615385 | 0.01 |
| 2188-10216 | 1-3 | 1. Male | 75.18891144 | 5 | 65.1162791 | 2.3355814 |
| 2209-10263 | 1-3 | 1. Male | 3829.77124 | 5 | 29.7297297 | 2.7127027 |
| 2228-10360 | 4-5 | 1. Male | 137.6486816 | 5 | 55.1724138 | 17.2513793 |
| 2233-10384 | 4-5 | 2. Female | 8689.171875 | 5 | 55.1724138 | 17.2513793 |
| 2243-10433 | 1-3 | 2. Female | 95.95072937 | 5 | 25 | 0.01 |
| 2243-10434 | 1-3 | 1. Male | 143.1248932 | 5 | 75.6756757 | 51.3613514 |
| 2243-10436 | 1-3 | 1. Male | 14.89410591 | 5 | 75.6756757 | 51.3613514 |
| 2244-10438 | 1-3 | 1. Male | 237.6708374 | 5 | 75.6756757 | 51.3613514 |
| 2246-10448 | 1-3 | 2. Female | 17.24055099 | 5 | 37.037037 | 11.1211111 |
| 2247-10454 | 1-3 | 1. Male | 29.19330406 | 5 | 75.6756757 | 51.3613514 |
| 2247-10456 | 1-3 | 1. Male | 80.50160217 | 5 | 75.6756757 | 51.3613514 |
| 2248-10461 | 4-5 | 2. Female | 72.31204987 | 5 | 63.4146342 | 4.88804878 |
| 2250-10467 | 1-3 | 2. Female | 226.358551 | 5 | 75.6756757 | 51.3613514 |
| 2251-10472 | 1-3 | 2. Female | 7149.289551 | 5 | 75.6756757 | 51.3613514 |
| 2253-10484 | 4-5 | 2. Female | 8.795941353 | 5 | 75.6756757 | 51.3613514 |
| 2253-10486 | 1-3 | 1. Male | 18.10214043 | 5 | 75.6756757 | 51.3613514 |
| 2254-10487 | 1-3 | 2. Female | 22.00116348 | 5 | 37.037037 | 11.1211111 |
| 2255-10493 | 4-5 | 1. Male | 9.235519409 | 5 | 75.6756757 | 51.3613514 |
| 2255-10495 | 1-3 | 1. Male | 26.73998451 | 5 | 75.6756757 | 51.3613514 |
| 2255-10496 | 1-3 | 2. Female | 2.728907824 | 5 | 75.6756757 | 51.3613514 |
| 2256-10501 | 1-3 | 2. Female | 922.0392456 | 5 | 75.6756757 | 51.3613514 |
| 2258-10504 | 1-3 | 2. Female | 46.1705246 | 5 | 46.3414634 | 26.8392683 |
| 2262-10525 | 4-5 | 2. Female | 9.984987259 | 5 | 43.75 | 6.26 |
| 2266-10545 | 1-3 | 1. Male | 913.0899048 | 5 | 50 | 5.01 |
| 2267-10551 | 1-3 | 1. Male | 11.44585896 | 5 | 50 | 5.01 |
| 2268-10556 | 1-3 | 2. Female | 1088.321655 | 5 | 21.2121212 | 3.04030303 |
| 2280-10615 | 1-3 | 1. Male | 35.13687134 | 5 | 46.3414634 | 26.8392683 |
| 2284-10629 | 4-5 | 2. Female | 4.995824337 | 5 | 53.4883721 | 41.8704651 |
| 2289-10657 | 4-5 | 1. Male | 20.34976768 | 5 | 46.875 | 9.385 |
| 2290-10659 | 4-5 | 2. Female | 20.1522522 | 5 | 60.6060606 | 18.1918182 |
| 2290-10661 | NA | NA | 3.801939964 | 5 | 60.6060606 | 18.1918182 |
| 2293-10676 | 4-5 | 2. Female | 69.5452652 | 5 | 53.4883721 | 41.8704651 |
| 2294-10681 | 4-5 | 1. Male | 120.0802002 | 5 | 53.4883721 | 41.8704651 |
| 2297-10696 | 4-5 | 1. Male | 3.692308903 | 5 | 46.875 | 9.385 |

| | | | | | | |
|---|---|---|---|---|---|---|
| 2300-10712 | 4-5 | 1. Male | 3.349193573 | 5 | 59.375 | 0.01 |
| 2303-10728 | 4-5 | 1. Male | 80.50160217 | 5 | 46.875 | 9.385 |
| 2311-10767 | 1-3 | 2. Female | 101.7332993 | 5 | 53.4883721 | 41.8704651 |
| 2314-10783 | 4-5 | 2. Female | 272.4438171 | 5 | 53.4883721 | 41.8704651 |
| 2316-10792 | 1-3 | 1. Male | 4.315890789 | 5 | 53.4883721 | 41.8704651 |
| 2316-10793 | 1-3 | 2. Female | 16.90750885 | 5 | 46.875 | 9.385 |
| 2356-10989 | 4-5 | 1. Male | 426.6997986 | 5 | 37.5 | 5.01 |
| 2394-11155 | 1-3 | 2. Female | 21.36673737 | 5 | 51.3513514 | 21.6316216 |
| 2405-11212 | 4-5 | 2. Female | 230.8174744 | 5 | 51.3513514 | 21.6316216 |
| 2437-11365 | NA | NA | 715.5155029 | 5 | 58.3333333 | 5.56555556 |
| 2437-11366 | 1-3 | 2. Female | 1444.092041 | 5 | 52.631579 | 26.3257895 |
| 2441-11387 | 1-3 | 1. Male | 240.0002899 | 5 | 58.3333333 | 5.56555556 |
| 2442-11393 | 4-5 | 1. Male | 141.7358398 | 5 | 17.6470588 | 2.95117647 |
| 2448-11422 | 1-3 | 2. Female | 197.4676056 | 5 | 33.3333333 | 11.1211111 |
| 2450-11434 | 1-3 | 2. Female | 16.41996384 | 5 | 33.3333333 | 11.1211111 |
| 2451-11436 | 4-5 | 2. Female | 27.26670837 | 5 | 33.3333333 | 11.1211111 |
| 2452-11442 | 4-5 | 1. Male | 82.08728027 | 5 | 17.6470588 | 2.95117647 |
| 2490-11623 | 1-3 | 1. Male | 4.71186161 | 5 | 38.0952381 | 35.7242857 |
| 2495-11650 | 4-5 | 2. Female | 484.3815613 | 5 | 38.0952381 | 35.7242857 |
| 2499-11670 | 4-5 | 1. Male | 6.252160072 | 5 | 40.8163265 | 18.3773469 |
| 2934-13737 | 1-3 | 2. Female | 15.04008579 | 3 | 59.1836735 | 4.09163265 |
| 2971-13918 | 1-3 | 2. Female | 6.437797546 | 3 | 70.212766 | 2.13765957 |
| 2976-13947 | 1-3 | 2. Female | 25.71686745 | 3 | 63.0434783 | 17.4013044 |
| 2977-13949 | 1-3 | 1. Male | 14.74954987 | 3 | 46.4285714 | 8.93857143 |
| 2983-13981 | 1-3 | 2. Female | 372.2389221 | 3 | 70.212766 | 2.13765957 |
| 2987-13999 | 1-3 | 2. Female | 612.1341553 | 3 | 70.212766 | 2.13765957 |
| 2991-14016 | 1-3 | 2. Female | 1430.076904 | 3 | 16.2790698 | 9.31232558 |
| 2994-14032 | 1-3 | 1. Male | 8604.842773 | 3 | 71.4285714 | 23.8195238 |
| 3001-14067 | 1-3 | 1. Male | 14.74954987 | 3 | 58.8235294 | 13.7354902 |
| 3002-14069 | 1-3 | 1. Male | 344.2988586 | 3 | 71.4285714 | 23.8195238 |
| 3003-14078 | 4-5 | 2. Female | 224.1615143 | 3 | 58.8235294 | 13.7354902 |
| 3004-14083 | 4-5 | 2. Female | 56.66516876 | 3 | 67.3913044 | 4.35782609 |
| 3027-14195 | 4-5 | 2. Female | 64.95569611 | 3 | 71.4285714 | 23.8195238 |
| 3029-14204 | 1-3 | 1. Male | 52795.60547 | 3 | 71.4285714 | 23.8195238 |
| 32-154 | 1-3 | 2. Female | 24.49284744 | 4 | 69.3548387 | 8.07451613 |
| 3327-15612 | 4-5 | 1. Male | 28.35148621 | 7 | 32.7 | 2.01 |
| 3334-15649 | 4-5 | 2. Female | 7.166893482 | 7 | 47.4 | 21.11 |
| 3335-15652 | 4-5 | 2. Female | 6.564609051 | 7 | 47.4 | 21.11 |
| 3380-15877 | 4-5 | 1. Male | 3.656471014 | 7 | 47.4 | 21.11 |
| 3387-15914 | 1-3 | 1. Male | 1.977916598 | 7 | 47.4 | 21.11 |
| 3395-15950 | 4-5 | 1. Male | 164.0651093 | 7 | 50.9 | 24.51 |
| 3396-15955 | 1-3 | 2. Female | 4.804675579 | 7 | 64 | 10.01 |
| 3399-15973 | 4-5 | 2. Female | 113.2547836 | 7 | 50.9 | 24.51 |
| 3417-16061 | 4-5 | 2. Female | 10.69049644 | 7 | 55.6 | 5.61 |
| 3433-16143 | 1-3 | 1. Male | 17.24055099 | 7 | 40.4 | 4.31 |
| 3437-16160 | 4-5 | 1. Male | 2.451292992 | 7 | 79.1 | 16.31 |

| | | | | | | |
|---|---|---|---|---|---|---|
| 3438-16167 | 1-3 | 1. Male | 68.87025452 | 7 | 57.1 | 2.41 |
| 3447-16212 | 4-5 | 2. Female | 24.7328949 | 7 | 39 | 9.81 |
| 3454-16248 | 4-5 | 2. Female | 5.14415884 | 7 | 50.9 | 24.51 |
| 3457-16264 | 1-3 | 2. Female | 10.90107727 | 7 | 56.4 | 10.91 |
| 3458-16265 | 4-5 | 2. Female | 51.39946747 | 7 | 55.6 | 5.61 |
| 3458-16266 | 4-5 | 2. Female | 27.53395462 | 7 | 50.9 | 24.51 |
| 3459-16273 | 4-5 | 1. Male | 102.7304077 | 7 | 57.1 | 2.41 |
| 3462-16289 | 1-3 | 2. Female | 19.38119125 | 7 | 39.7 | 12.11 |
| 3508-16516 | 1-3 | 2. Female | 36.18014526 | 7 | 66.7 | 9.81 |
| 3520-16577 | 1-3 | 1. Male | 116.6175156 | 7 | 50 | 11.91 |
| 3532-16637 | 4-5 | 2. Female | 280.5331421 | 7 | 50 | 11.91 |
| 3538-16667 | 1-3 | 2. Female | 138.9978027 | 7 | 43.9 | 17.11 |
| 3543-16689 | 1-3 | 1. Male | 175225.9688 | 7 | 61.5 | 7.71 |
| 3544-16697 | 1-3 | 1. Male | 84.52458954 | 7 | 54.7 | 4.71 |
| 3566-16805 | 1-3 | 1. Male | 39788.73828 | 7 | 37.5 | 2.11 |
| 3573-16841 | 4-5 | 2. Female | 31.25602722 | 7 | 54.7 | 4.71 |
| 3773-17820 | 1-3 | 2. Female | 25.96892357 | 9 | 47.6 | 11.91 |
| 3778-17846 | 4-5 | 1. Male | 36.89281845 | 9 | 47.6 | 11.91 |
| 3780-17855 | 1-3 | 2. Female | 49.91728973 | 9 | 47.6 | 11.91 |
| 3780-17857 | 1-3 | 1. Male | 13.24906158 | 9 | 53.1 | 3.11 |
| 3793-17922 | 4-5 | 2. Female | 655.3856812 | 9 | 27.9 | 2.31 |
| 3797-17943 | 1-3 | 2. Female | 6.693918705 | 9 | 27.9 | 2.31 |
| 3811-18007 | 4-5 | 1. Male | 92.27948761 | 9 | 41.3 | 0.01 |
| 3816-18033 | 4-5 | 2. Female | 170.5922546 | 9 | 41.3 | 0.01 |
| 3822-18059 | 1-3 | 2. Female | 19.76296043 | 9 | 44.6 | 10.71 |
| 3839-18146 | 1-3 | 1. Male | 1.847386003 | 9 | 42.6 | 5.61 |
| 42-207 | 4-5 | 2. Female | 84.52458954 | 4 | 69.3548387 | 8.07451613 |
| 45-216 | 4-5 | 1. Male | 1576.583496 | 4 | 90.6976744 | 39.5448837 |
| 45-218 | 4-5 | 1. Male | 141.7358398 | 4 | 46.1538462 | 3.85615385 |
| 48-231 | 1-3 | 2. Female | 20.75061607 | 4 | 90.6976744 | 39.5448837 |
| 5383-36710 | 1-3 | 1. Male | 4.758040905 | 13 | 61.3 | 16.11 |
| 5386-36723 | 4-5 | 2. Female | 303.298584 | 13 | 38.1 | 4.81 |
| 5389-36737 | 4-5 | 2. Female | 30.65224075 | 13 | 38.1 | 4.81 |
| 5390-36742 | 4-5 | 1. Male | 28.90995216 | 13 | 38.1 | 4.81 |
| 5392-36753 | 4-5 | 2. Female | 80.50160217 | 13 | 55 | 25.01 |
| 5395-36766 | 1-3 | 2. Female | 357.9963989 | 13 | 38.1 | 4.81 |
| 5395-36770 | 1-3 | 2. Female | 39.88671494 | 13 | 55 | 25.01 |
| 64-311 | 4-5 | 2. Female | 24.97530746 | 4 | 55.1724138 | 31.0444828 |
| 64-314 | 1-3 | 1. Male | 6.437797546 | 4 | 90.6976744 | 39.5448837 |
| 65-316 | 1-3 | 1. Male | 88.74871063 | 4 | 58.0645161 | 22.5906452 |
| 70-340 | 4-5 | 2. Female | 12.74212933 | 4 | 55.1724138 | 31.0444828 |
| 71-347 | 1-3 | 2. Female | 5.782876015 | 4 | 60.6060606 | 12.1312121 |
| 73-355 | 1-3 | 1. Male | 70.22689056 | 4 | 64.9122807 | 21.0626316 |
| 75-367 | 1-3 | 2. Female | 2971.959473 | 4 | 78.2608696 | 13.0534783 |
| 77-379 | 1-3 | 1. Male | 19.76296043 | 4 | 64.9122807 | 21.0626316 |
| 78-382 | 4-5 | 1. Male | 6.960232258 | 4 | 58.0645161 | 22.5906452 |

| | | | | | | |
|---|---|---|---|---|---|---|
| 850-3968 | 1-3 | 2. Female | 758.6360474 | 6 | 79.3103448 | 3.45827586 |
| 859-3999 | 1-3 | 1. Male | 513.5733643 | 6 | 71.1538462 | 17.3176923 |
| 865-4011 | 1-3 | 1. Male | 107.8642578 | 6 | 71.1538462 | 17.3176923 |
| 866-4014 | 4-5 | 2. Female | 668.295166 | 6 | 81.4814815 | 11.1211111 |
| 868-4024 | 1-3 | 1. Male | 4.031065941 | 6 | 83.7209302 | 18.6146512 |
| 872-4044 | 1-3 | 1. Male | 111.0670395 | 6 | 56.4516129 | 27.4293548 |
| 875-4061 | 1-3 | 1. Male | 4.35819149 | 6 | 83.7209302 | 18.6146512 |
| 883-4100 | 1-3 | 2. Female | 12.74212933 | 6 | 83.7209302 | 18.6146512 |
| 884-4103 | 1-3 | 2. Female | 58.91953278 | 6 | 83.7209302 | 18.6146512 |
| 889-4129 | 1-3 | 2. Female | 85.35303497 | 6 | 83.7209302 | 18.6146512 |
| 889-4130 | 4-5 | 2. Female | 39.11618805 | 6 | 83.7209302 | 18.6146512 |

| tf_dv | ti_dv | ts_dv | porf2_ul | omcb_ul | TF_Dist | TI_Dist |
|---|---|---|---|---|---|---|
| 1. Yes | 0. No | 0. No | 201.300355 | 46.8385704 | 17.8 | 4.3 |
| 0. No | 0. No | 0. No | 120.280487 | 28.7642294 | 18.8 | 6.2 |
| 1. Yes | 0. No | 0. No | 53.3908332 | 28.8505159 | 18.8 | 6.2 |
| 0. No | 0. No | 0. No | 12.0265161 | 6.15848525 | 18.8 | 6.2 |
| 0. No | 0. No | 0. No | NA | NA | 18.8 | 6.2 |
| 0. No | 1. Yes | 0. No | NA | NA | 18.8 | 6.2 |
| 0. No | 0. No | 1. Yes | 34.1504944 | 11.6627178 | 24.3 | 6.5 |
| 1. Yes | 1. Yes | 0. No | 83.3355826 | 27.7061638 | 23.9 | 8.8 |
| 0. No | 0. No | 0. No | NA | 297.280928 | 23.9 | 8.8 |
| 1. Yes | 0. No | 0. No | 41.2820224 | 9.78043356 | 23.9 | 8.8 |
| 1. Yes | 1. Yes | 0. No | 433.815841 | 113.510589 | 23.9 | 8.8 |
| 1. Yes | 1. Yes | 0. No | NA | 93.6136872 | 23.9 | 8.8 |
| 0. No | 0. No | 0. No | 228.749135 | 71.454164 | 23.9 | 8.8 |
| 1. Yes | 0. No | 0. No | 386.693194 | 67.4011162 | 23.9 | 8.8 |
| 0. No | 0. No | 0. No | NA | 135.054936 | 23.9 | 8.8 |
| 1. Yes | 1. Yes | 0. No | NA | NA | 23.9 | 8.8 |
| 1. Yes | 1. Yes | 0. No | NA | NA | 23.9 | 8.8 |
| 1. Yes | 0. No | 0. No | NA | NA | 23.9 | 8.8 |
| 0. No | 1. Yes | 0. No | 247.735204 | 53.7087063 | 23.9 | 8.8 |
| 1. Yes | 0. No | 0. No | 211.388326 | 50.6027708 | 33.8 | 9.7 |
| 1. Yes | 0. No | 0. No | 71.5939156 | 25.1466777 | 33.8 | 9.7 |
| 1. Yes | 1. Yes | 0. No | 136.218761 | 42.532198 | 33.8 | 9.7 |
| 1. Yes | 1. Yes | 0. No | 119.303071 | 29.4769413 | 33.8 | 9.7 |
| 1. Yes | 0. No | 0. No | 291.571188 | 68.1090396 | 33.8 | 9.7 |
| 1. Yes | 0. No | 0. No | NA | NA | 33.8 | 9.7 |
| 1. Yes | 1. Yes | 0. No | NA | NA | 33.8 | 9.7 |
| 1. Yes | 1. Yes | 0. No | 72.8215031 | 18.9221291 | 44.9 | 10.7 |
| 1. Yes | 0. No | 0. No | 254.159874 | 63.8600818 | 44.9 | 10.7 |
| 1. Yes | 1. Yes | 0. No | NA | 2028.7985 | 44.9 | 10.7 |
| 0. No | 1. Yes | 0. No | 317.379283 | 68.9219974 | 44.9 | 10.7 |
| 1. Yes | 0. No | 0. No | 288.739021 | 25.4946442 | 44.9 | 10.7 |
| 1. Yes | 1. Yes | 0. No | 79.0142154 | 31.1654079 | 30.6 | 9.5 |
| 1. Yes | 1. Yes | 0. No | 86.308474 | 28.4289973 | 30.6 | 9.5 |
| 0. No | 0. No | 0. No | 98.2225066 | 15.7925791 | 30.6 | 9.5 |
| 1. Yes | 1. Yes | 0. No | 133.646009 | 35.1027678 | 30.6 | 9.5 |
| 1. Yes | 1. Yes | 0. No | NA | NA | 30.6 | 9.5 |
| 0. No | 0. No | 0. No | NA | NA | 30.6 | 9.5 |
| 1. Yes | 1. Yes | 0. No | NA | NA | 30.6 | 9.5 |
| 1. Yes | 1. Yes | 0. No | 261.193 | 70.3935279 | 30.6 | 9.5 |
| 1. Yes | 1. Yes | 0. No | 1315.81524 | 485.07641 | 51.8 | 16.3 |
| 1. Yes | 0. No | 0. No | 893.372126 | 141.033177 | 51.8 | 16.3 |
| 1. Yes | 1. Yes | 0. No | 1145.18019 | 237.523296 | 51.8 | 16.3 |
| 1. Yes | 0. No | 0. No | 1990.16478 | 160.821421 | 51.8 | 16.3 |
| 1. Yes | 0. No | 0. No | NA | NA | 51.8 | 16.3 |
| 1. Yes | 0. No | 0. No | NA | NA | 36.1 | 8.2 |

| | | | | | | |
|---|---|---|---|---|---|---|
| 0. No | 1. Yes | 0. No | 1574.64062 | 428.416363 | 36.1 | 8.2 |
| 1. Yes | 0. No | 0. No | 444.350244 | 180.393583 | 36.1 | 8.2 |
| 1. Yes | 0. No | 0. No | NA | NA | 36.1 | 8.2 |
| 0. No | 0. No | 0. No | NA | 1526.55315 | 48.1 | 6.5 |
| 1. Yes | 0. No | 0. No | NA | NA | 48.1 | 6.5 |
| 1. Yes | 1. Yes | 0. No | NA | 170.528944 | 48.1 | 6.5 |
| 1. Yes | 0. No | 0. No | NA | NA | 48.1 | 6.5 |
| 1. Yes | 0. No | 0. No | NA | NA | 48.1 | 6.5 |
| 1. Yes | 0. No | 0. No | NA | NA | 48.1 | 6.5 |
| 1. Yes | 0. No | 0. No | NA | 1716.34048 | 48.1 | 6.5 |
| 1. Yes | 0. No | 0. No | 152.686839 | 39.9758134 | 48.1 | 6.5 |
| 0. No | 0. No | 0. No | 636.221628 | 357.561058 | 48.1 | 6.5 |
| 1. Yes | 0. No | 0. No | 160.419434 | 73.9853216 | 48.1 | 6.5 |
| 0. No | 1. Yes | 0. No | NA | 1821.75952 | 39.2 | 3.5 |
| 1. Yes | 1. Yes | 0. No | 518.296133 | 171.599534 | 28.5 | 5 |
| 1. Yes | 1. Yes | 0. No | NA | 471.059518 | 28.5 | 5 |
| 1. Yes | 0. No | 0. No | 576.998142 | 230.541361 | 42.4 | 13.8 |
| 1. Yes | 1. Yes | 0. No | 443.353361 | 163.962117 | 42.4 | 13.8 |
| 1. Yes | 1. Yes | 0. No | NA | NA | 42.4 | 13.8 |
| 1. Yes | 1. Yes | 0. No | 634.732474 | 221.372253 | 42.4 | 13.8 |
| 1. Yes | 0. No | 0. No | NA | NA | 42.4 | 13.8 |
| 1. Yes | 1. Yes | 0. No | 409.371105 | 130.857149 | 42.4 | 13.8 |
| 1. Yes | 0. No | 0. No | 363.048082 | 118.217785 | 42.4 | 13.8 |
| 1. Yes | 0. No | 0. No | 517.786995 | 154.665644 | 42.4 | 13.8 |
| 1. Yes | 1. Yes | 0. No | 740.855257 | 245.813544 | 42.4 | 13.8 |
| 1. Yes | 1. Yes | 0. No | 5536.18289 | 1481.66155 | 42.4 | 13.8 |
| 1. Yes | 1. Yes | 0. No | NA | NA | 42.4 | 13.8 |
| 1. Yes | 1. Yes | 0. No | NA | NA | 42.4 | 13.8 |
| 0. No | 1. Yes | 0. No | 217.054233 | 56.7752371 | 42.4 | 13.8 |
| 1. Yes | 1. Yes | 0. No | NA | NA | 42.4 | 13.8 |
| 1. Yes | 0. No | 0. No | 419.470055 | 388.309917 | 42.4 | 13.8 |
| 1. Yes | 0. No | 0. No | NA | NA | 42.4 | 13.8 |
| 1. Yes | 1. Yes | 0. No | 1141.90267 | 331.630714 | 42.4 | 13.8 |
| 1. Yes | 1. Yes | 0. No | 335.521697 | 143.144842 | 34.6 | 10.1 |
| 0. No | 0. No | 0. No | NA | NA | 34.6 | 10.1 |
| 1. Yes | 0. No | 0. No | 1775.93544 | 806.058249 | 34.6 | 10.1 |
| 1. Yes | 0. No | 0. No | NA | NA | 34.6 | 10.1 |
| 0. No | 1. Yes | 0. No | 2060.61083 | 805.152598 | 34.6 | 10.1 |
| 1. Yes | 1. Yes | 0. No | 508.030174 | 145.459044 | 34.6 | 10.1 |
| 1. Yes | 1. Yes | 0. No | NA | NA | 41.8 | 11.6 |
| 1. Yes | 0. No | 0. No | NA | NA | 41.8 | 11.6 |
| 1. Yes | 0. No | 0. No | NA | NA | 41.8 | 11.6 |
| NA | NA | NA | NA | NA | 41.8 | 11.6 |
| 1. Yes | 1. Yes | 0. No | 588.193841 | 220.349677 | 41.8 | 11.6 |
| 1. Yes | 1. Yes | 0. No | 651.892878 | 156.671921 | 41.8 | 11.6 |
| 1. Yes | 0. No | 0. No | NA | NA | 41.8 | 11.6 |

| | | | | | | |
|---|---|---|---|---|---|---|
| 1. Yes | 0. No | 0. No | 13.7142283 | 7.14273723 | 41.8 | 11.6 |
| 1. Yes | 0. No | 0. No | 860.801698 | 218.740927 | 41.8 | 11.6 |
| 1. Yes | 0. No | 0. No | 1.53089755 | 1.12985593 | 41.8 | 11.6 |
| 1. Yes | 1. Yes | 0. No | 783.900539 | 245.505197 | 41.8 | 11.6 |
| 1. Yes | 1. Yes | 0. No | NA | NA | 41.8 | 11.6 |
| 1. Yes | 0. No | 0. No | NA | NA | 41.8 | 11.6 |
| 1. Yes | 1. Yes | 0. No | 1105.15283 | 202.497681 | 35.8 | 4.9 |
| 1. Yes | 1. Yes | 0. No | 172.186105 | 48.1608956 | 33.8 | 5.9 |
| 1. Yes | 0. No | 0. No | 855.50826 | 90.917979 | 33.8 | 5.9 |
| NA | NA | NA | 1256.2074 | 456.685001 | 39.8 | 9.6 |
| 0. No | 1. Yes | 0. No | 2610.35488 | 1402.22543 | 39.8 | 9.6 |
| 0. No | 1. Yes | 0. No | 1095.27845 | 437.860984 | 39.8 | 9.6 |
| 1. Yes | 0. No | 0. No | 1104.0834 | 427.31236 | 39.8 | 9.6 |
| 1. Yes | 0. No | 0. No | 1622.95178 | 703.413885 | 39.8 | 9.6 |
| 0. No | 1. Yes | 0. No | NA | NA | 39.8 | 9.6 |
| 0. No | 0. No | 0. No | 108.782674 | 48.4725219 | 39.8 | 9.6 |
| 1. Yes | 0. No | 0. No | 176.0866 | 57.4189874 | 39.8 | 9.6 |
| 1. Yes | 1. Yes | 0. No | NA | NA | 33.8 | 9.7 |
| 0. No | 1. Yes | 0. No | 636.307801 | 184.440494 | 33.8 | 9.7 |
| 1. Yes | 0. No | 0. No | NA | NA | 33.8 | 9.7 |
| 1. Yes | 0. No | 0. No | NA | NA | 24.5 | 3.7 |
| 1. Yes | 0. No | 0. No | NA | NA | 41.1 | 7.7 |
| 1. Yes | 1. Yes | 0. No | 264.739619 | 51.5268816 | 41.1 | 7.7 |
| 0. No | 1. Yes | 0. No | NA | NA | 41.1 | 7.7 |
| 1. Yes | 0. No | 0. No | 243.917988 | 77.5971613 | 41.1 | 7.7 |
| 1. Yes | 0. No | 0. No | 155.358239 | 64.5400135 | 41.1 | 7.7 |
| 1. Yes | 0. No | 0. No | 339.440465 | 103.123396 | 27.6 | 7.8 |
| 1. Yes | 1. Yes | 0. No | NA | 115.815582 | 27.6 | 7.8 |
| 1. Yes | 0. No | 0. No | NA | NA | 27.6 | 7.8 |
| 1. Yes | 0. No | 0. No | 238.707766 | 88.3749311 | 27.6 | 7.8 |
| 0. No | 1. Yes | 0. No | 221.157189 | 82.9690385 | 27.6 | 7.8 |
| 1. Yes | 0. No | 0. No | 80.7635691 | 16.6583867 | 27.6 | 7.8 |
| 1. Yes | 0. No | 0. No | 83.9912562 | 21.031085 | 27.6 | 7.8 |
| 1. Yes | 1. Yes | 0. No | NA | 1300.64415 | 27.6 | 7.8 |
| 1. Yes | 0. No | 0. No | 184.086146 | 46.5618139 | 73.9 | 8 |
| 1. Yes | 0. No | 0. No | 174.351208 | 44.236064 | 29.1 | 5 |
| 1. Yes | 1. Yes | 0. No | NA | NA | 29.1 | 5 |
| 1. Yes | 0. No | 0. No | NA | NA | 29.1 | 5 |
| 1. Yes | 0. No | 0. No | NA | NA | 29.1 | 5 |
| 0. No | 1. Yes | 0. No | NA | NA | 29.1 | 5 |
| 1. Yes | 0. No | 0. No | 333.175224 | 120.75662 | 49.5 | 9.5 |
| 1. Yes | 1. Yes | 1. Yes | NA | NA | 49.5 | 9.5 |
| 1. Yes | 0. No | 0. No | 340.757429 | 147.378736 | 49.5 | 9.5 |
| 1. Yes | 1. Yes | 0. No | NA | NA | 49.5 | 9.5 |
| 1. Yes | 0. No | 0. No | NA | NA | 49.5 | 9.5 |
| 1. Yes | 1. Yes | 0. No | NA | NA | 49.5 | 9.5 |

| | | | | | | |
|---|---|---|---|---|---|---|
| 1. Yes | 0. No | 0. No | 165.339724 | 51.2749192 | 49.5 | 9.5 |
| 1. Yes | 1. Yes | 0. No | 321.090671 | 41.9051725 | 49.5 | 9.5 |
| 0. No | 1. Yes | 0. No | NA | NA | 49.5 | 9.5 |
| 0. No | 1. Yes | 0. No | NA | NA | 49.5 | 9.5 |
| 1. Yes | 0. No | 0. No | 142.391471 | 38.1352591 | 49.5 | 9.5 |
| 1. Yes | 1. Yes | 0. No | 80.6910744 | 32.373195 | 49.5 | 9.5 |
| 1. Yes | 0. No | 0. No | 522.038059 | 189.899674 | 49.5 | 9.5 |
| 1. Yes | 0. No | 0. No | NA | NA | 35.8 | 7.3 |
| 1. Yes | 1. Yes | 0. No | 188.890468 | 53.9064127 | 35.8 | 7.3 |
| 1. Yes | 1. Yes | 0. No | 775.446372 | 254.891363 | 35.8 | 7.4 |
| 1. Yes | 0. No | 0. No | 668.682842 | 230.971083 | 35.8 | 7.4 |
| 1. Yes | 1. Yes | 0. No | 278.232786 | 64.1587688 | 35.8 | 7.4 |
| 1. Yes | 0. No | 0. No | NA | 18042.2086 | 35.6 | 4.9 |
| 1. Yes | 0. No | 0. No | 143.169667 | 52.9249419 | 35.6 | 4.9 |
| 0. No | 0. No | 0. No | NA | 1219.20358 | 35.6 | 4.9 |
| 0. No | 0. No | 0. No | 123.942589 | 20.1812657 | 35.6 | 4.9 |
| 1. Yes | 1. Yes | 0. No | 390.667106 | 225.605518 | 32.6 | 6.7 |
| 1. Yes | 0. No | 0. No | 206.738801 | 103.472168 | 32.6 | 6.7 |
| 1. Yes | 1. Yes | 0. No | 463.294604 | 167.001394 | 32.6 | 6.7 |
| 1. Yes | 0. No | 0. No | NA | NA | 32.6 | 6.7 |
| 0. No | 0. No | 0. No | 464.702713 | 126.894028 | 20.7 | 3.7 |
| 1. Yes | 0. No | 0. No | NA | NA | 20.7 | 3.7 |
| 1. Yes | 0. No | 0. No | NA | NA | 47.8 | 9.6 |
| 1. Yes | 0. No | 0. No | 460.146968 | 141.220976 | 47.8 | 9.6 |
| 1. Yes | 1. Yes | 0. No | NA | NA | 45.7 | 8.4 |
| 1. Yes | 1. Yes | 0. No | NA | NA | 45.7 | 8.4 |
| 1. Yes | 0. No | 0. No | 452.18961 | 137.356458 | 73.9 | 8 |
| 1. Yes | 1. Yes | 0. No | 1388.33755 | 287.065513 | 62 | 21.4 |
| 1. Yes | 0. No | 0. No | 2954.90642 | 1122.00055 | 62 | 21.4 |
| 1. Yes | 1. Yes | 0. No | NA | NA | 62 | 21.4 |
| 1. Yes | 0. No | NA | NA | NA | 36.96 | 6.2 |
| 1. Yes | 0. No | NA | 201.277466 | 62.8366808 | 36.96 | 6.2 |
| 1. Yes | 0. No | NA | 232.454556 | 41.5817723 | 36.96 | 6.2 |
| 1. Yes | 0. No | NA | 173.311427 | 28.7748288 | 36.96 | 6.2 |
| 1. Yes | 1. Yes | NA | 122.363369 | 48.6953082 | 36.96 | 6.2 |
| 1. Yes | 0. No | NA | NA | NA | 36.96 | 6.2 |
| 1. Yes | 0. No | NA | 248.637628 | 68.757835 | 36.96 | 6.2 |
| 1. Yes | 1. Yes | 0. No | 429.926871 | 125.13909 | 62 | 21.4 |
| 1. Yes | 0. No | 0. No | NA | NA | 62 | 21.4 |
| 1. Yes | 1. Yes | 0. No | 1775.5108 | 530.872465 | 62 | 21.4 |
| 1. Yes | 1. Yes | 0. No | NA | NA | 62 | 21.4 |
| 1. Yes | 0. No | 0. No | NA | NA | 62 | 21.4 |
| 0. No | 1. Yes | 0. No | 888.35864 | 407.634929 | 62 | 21.4 |
| 1. Yes | 0. No | 0. No | NA | 1145.63417 | 62 | 21.4 |
| 1. Yes | 1. Yes | 0. No | NA | NA | 62 | 21.4 |
| 1. Yes | 0. No | 0. No | NA | NA | 62 | 21.4 |

| | | | | | | |
|---|---|---|---|---|---|---|
| 1. Yes | 0. No | 0. No | 443.919333 | 188.219808 | 58.9 | 13.4 |
| 1. Yes | 0. No | 0. No | 274.338636 | 49.5509187 | 65.3 | 13.9 |
| 1. Yes | 1. Yes | 0. No | 249.561181 | 77.3758922 | 65.3 | 13.9 |
| 1. Yes | 0. No | 0. No | 197.335206 | 45.2463667 | 55.6 | 14.2 |
| 1. Yes | 0. No | 0. No | NA | NA | 55.6 | 14.2 |
| 1. Yes | 1. Yes | 0. No | 127.900907 | 39.3536285 | 55.6 | 14.2 |
| 1. Yes | 1. Yes | 0. No | NA | NA | 55.6 | 14.2 |
| 1. Yes | 0. No | 0. No | NA | NA | 55.6 | 14.2 |
| 1. Yes | 0. No | 0. No | NA | 28.8970678 | 55.6 | 14.2 |
| 1. Yes | 1. Yes | 0. No | 645.178304 | 294.574024 | 55.6 | 14.2 |
| 1. Yes | 0. No | 0. No | 313.668433 | 132.871053 | 55.6 | 14.2 |

| Ct_Dist | MDA | CT1335_FLAN | CT1335 | ct1335_flank_comb. | CT1299_FLAN |
|---|---|---|---|---|---|
| 0.8 | ROUNDS_MDA5 | GAAAAGG | 10T/8A | GAAAAGG_10T/8A | ATTCT |
| 6.88 | ROUNDS_MDA5 | GAAAAGG | 10T/8A | GAAAAGG_10T/8A | ATTCT |
| 6.88 | ROUNDS_MDA5 | GAAAAGG | 10T/8A | GAAAAGG_10T/8A | ATTCT |
| 6.88 | ROUNDS_MDA5 | GAAAAGG | 10T/7A | GAAAAGG_10T/7A | ATTCT |
| 6.88 | ROUNDS_MDA5 | GAAAAAGG | 10T/8A | GAAAAAGG_10T/8A | ATTCT |
| 6.88 | ROUNDS_MDA5 | GAAAAAGG | 10T/8A | GAAAAAGG_10T/8A | ATTCT |
| 0.89 | ROUNDS_MDA5 | GAAAAGG | 10T/8A | GAAAAGG_10T/8A | ATTCT |
| 10.84 | ROUNDS_MDA5 | GAAAAGG | 10T/8A | GAAAAGG_10T/8A | ATTCT |
| 10.84 | ROUNDS_MDA5 | GAAAAGG | 10T/8A | GAAAAGG_10T/8A | ATTCT |
| 10.84 | ROUNDS_MDA5 | GAAAAAGG | 10T/7A | GAAAAAGG_10T/7A | ATTCT |
| 10.84 | ROUNDS_MDA5 | GAAAAGG | 10T/7A | GAAAAGG_10T/7A | ATTCT |
| 10.84 | ROUNDS_MDA5 | GAAAAAGG | 10T/8A | GAAAAAGG_10T/8A | ATTCT |
| 10.84 | ROUNDS_MDA5 | GAAAAGG | 10T/8A | GAAAAGG_10T/8A | ATTCT |
| 10.84 | ROUNDS_MDA5 | GAAAAGG | 10T/8A | GAAAAGG_10T/8A | ATTCT |
| 10.84 | ROUNDS_MDA5 | GAAAAGG | 10T/8A | GAAAAGG_10T/8A | ATTCT |
| 10.84 | ROUNDS_MDA5 | GAAAAAGG | 10T/7A | GAAAAAGG_10T/7A | ATTCT |
| 10.84 | ROUNDS_MDA5 | GAAAAGG | 10T/8A | GAAAAGG_10T/8A | ATTCT |
| 10.84 | ROUNDS_MDA5 | GAAAAGG | 10T/8A | GAAAAGG_10T/8A | ATTCT |
| 10.84 | ROUNDS_MDA5 | GAAAAGG | 10T/8A | GAAAAGG_10T/8A | ATTCT |
| 10.88 | ROUNDS_MDA5 | GAAAAGG | 10T/8A | GAAAAGG_10T/8A | ATTCT |
| 10.88 | ROUNDS_MDA5 | GAAAAGG | 10T/8A | GAAAAGG_10T/8A | ATTCT |
| 10.88 | ROUNDS_MDA5 | GAAAAGG | 10T/8A | GAAAAGG_10T/8A | ATTCT |
| 10.88 | ROUNDS_MDA5 | GAAAAAGG | 10T/8A | GAAAAAGG_10T/8A | ATTCT |
| 10.88 | ROUNDS_MDA5 | GAAAAAGG | 10T/8A | GAAAAAGG_10T/8A | ATTCT |
| 10.88 | ROUNDS_MDA5 | GAAAAGG | 10T/8A | GAAAAGG_10T/8A | ATTCT |
| 10.88 | ROUNDS_MDA5 | GAAAAGG | 10T/8A | GAAAAGG_10T/8A | ATTCT |
| 11.93 | ROUNDS_MDA5 | GAAAAGG | 10T/8A | GAAAAGG_10T/8A | ATTCT |
| 11.93 | ROUNDS_MDA5 | GAAAAGG | 10T/8A | GAAAAGG_10T/8A | ATTCT |
| 11.93 | ROUNDS_MDA5 | GAAAAGG | 10T/7A | GAAAAGG_10T/7A | ATTCT |
| 11.93 | ROUNDS_MDA5 | GAAAAGG | 10T/8A | GAAAAGG_10T/8A | ATTCT |
| 11.93 | ROUNDS_MDA5 | GAAAAGG | 10T/8A | GAAAAGG_10T/8A | ATTCT |
| 10.04 | ROUNDS_MDA5 | GAAAAAGG | 10T/7A | GAAAAAGG_10T/7A | ATTCT |
| 10.04 | ROUNDS_MDA5 | GAAAAGG | 10T/8A | GAAAAGG_10T/8A | ATTCT |
| 10.04 | ROUNDS_MDA5 | GAAAAGG | 10T/8A | GAAAAGG_10T/8A | ATTCT |
| 10.04 | ROUNDS_MDA5 | GAAAAGG | 10T/8A | GAAAAGG_10T/8A | ATTCT |
| 10.04 | ROUNDS_MDA5 | GAAAAGG | 10T/8A | GAAAAGG_10T/8A | ATTCT |
| 10.04 | ROUNDS_MDA5 | GAAAAGG | 10T/8A | GAAAAGG_10T/8A | ATTCT |
| 10.04 | ROUNDS_MDA5 | GAAAAGG | 10T/8A | GAAAAGG_10T/8A | ATTCT |
| 10.04 | ROUNDS_MDA5 | GAAAAGG | 10T/7A | GAAAAGG_10T/7A | ATTCT |
| 17.5 | ROUNDS_MDA5 | GAAAAGG | 10T/8A | GAAAAGG_10T/8A | ATTCT |
| 17.5 | ROUNDS_MDA5 | GAAAAGG | 10T/8A | GAAAAGG_10T/8A | ATTCT |
| 17.5 | ROUNDS_MDA5 | GAAAAGG | 10T/8A | GAAAAGG_10T/8A | ATTCT |
| 17.5 | ROUNDS_MDA5 | GAAAAGG | 10T/8A | GAAAAGG_10T/8A | ATTCT |
| 17.5 | ROUNDS_MDA5 | GAAAAAGG | 10T/8A | GAAAAAGG_10T/8A | ATTCT |
| 4.4 | ROUNDS_MDA5 | GAAAAGG | 10T/8A | GAAAAGG_10T/8A | ATTCT |

| | | | | | |
|---|---|---|---|---|---|
| 4.4 | ROUNDS_MDA5 | GAAAAGG | 10T/8A | GAAAAGG_10T/8A | ATTCT |
| 4.4 | ROUNDS_MDA5 | GAAAAGG | 10T/8A | GAAAAGG_10T/8A | ATTCT |
| 4.4 | ROUNDS_MDA5 | GAAAAAGG | 10T/8A | GAAAAAGG_10T/8A | ATTCT |
| 28.69 | ROUNDS_MDA5 | GAAAAGG | 10T/8A | GAAAAGG_10T/8A | ATTCT |
| 28.69 | ROUNDS_MDA5 | GAAAAAGG | 10T/8A | GAAAAAGG_10T/8A | ATTCT |
| 28.69 | ROUNDS_MDA5 | GAAAAGG | 10T/8A | GAAAAGG_10T/8A | ATTCT |
| 28.69 | ROUNDS_MDA5 | GAAAAAGG | 10T/7A | GAAAAAGG_10T/7A | ATTCT |
| 28.69 | ROUNDS_MDA5 | GAAAAAGG | 10T/8A | GAAAAAGG_10T/8A | ATTCT |
| 28.69 | ROUNDS_MDA5 | GAAAAAGG | 10T/8A | GAAAAAGG_10T/8A | ATTCT |
| 28.69 | ROUNDS_MDA5 | GAAAAGG | 10T/8A | GAAAAGG_10T/8A | ATTCT |
| 28.69 | ROUNDS_MDA5 | GAAAAGG | 9T/8A | GAAAAGG_9T/8A | ATTCT |
| 28.69 | ROUNDS_MDA5 | GAAAAGG | 10T/8A | GAAAAGG_10T/8A | ATTCT |
| 28.69 | ROUNDS_MDA5 | GAAAAGG | 10T/8A | GAAAAGG_10T/8A | ATTCT |
| 5.56 | ROUNDS_MDA5 | GAAAAGG | 9T/8A | GAAAAGG_9T/8A | ATTCT |
| 4.44 | ROUNDS_MDA5 | GAAAAGG | 10T/8A | GAAAAGG_10T/8A | ATTCT |
| 4.44 | ROUNDS_MDA5 | GAAAAGG | 10T/8A | GAAAAGG_10T/8A | ATTCT |
| 38.3 | ROUNDS_MDA5 | GAAAAAGG | 10T/8A | GAAAAAGG_10T/8A | ATTCT |
| 38.3 | ROUNDS_MDA5 | GAAAAGG | 10T/8A | GAAAAGG_10T/8A | ATTCT |
| 38.3 | ROUNDS_MDA5 | GAAAAGG | 10T/8A | GAAAAGG_10T/8A | ATTCT |
| 38.3 | ROUNDS_MDA5 | GAAAAAGG | 10T/7A | GAAAAAGG_10T/7A | ATTCT |
| 38.3 | ROUNDS_MDA5 | GAAAAGG | 10T/8A | GAAAAGG_10T/8A | ATTCT |
| 38.3 | ROUNDS_MDA5 | GAAAAGG | 10T/8A | GAAAAGG_10T/8A | ATTCT |
| 38.3 | ROUNDS_MDA5 | GAAAAGG | 10T/8A | GAAAAGG_10T/8A | ATTCT |
| 38.3 | ROUNDS_MDA5 | GAAAAGG | 10T/8A | GAAAAGG_10T/8A | ATTCT |
| 38.3 | ROUNDS_MDA5 | GAAAAAGG | 10T/7A | GAAAAAGG_10T/7A | ATTCT |
| 38.3 | ROUNDS_MDA5 | GAAAAAGG | 10T/8A | GAAAAAGG_10T/8A | ATTCT |
| 38.3 | ROUNDS_MDA5 | GAAAAAGG | 10T/8A | GAAAAAGG_10T/8A | ATTCT |
| 38.3 | ROUNDS_MDA5 | GAAAAAGG | 10T/7A | GAAAAAGG_10T/7A | ATTCT |
| 38.3 | ROUNDS_MDA5 | GAAAAGG | 10T/8A | GAAAAGG_10T/8A | ATTCT |
| 38.3 | ROUNDS_MDA5 | GAAAAAGG | 10T/8A | GAAAAAGG_10T/8A | ATTCT |
| 38.3 | ROUNDS_MDA5 | GAAAAGG | 10T/8A | GAAAAGG_10T/8A | ATTCT |
| 38.3 | ROUNDS_MDA5 | GAAAAGG | 10T/8A | GAAAAGG_10T/8A | ATTCT |
| 38.3 | ROUNDS_MDA5 | GAAAAGG | 10T/8A | GAAAAGG_10T/8A | ATTCT |
| 14.29 | ROUNDS_MDA5 | GAAAAGG | 10T/8A | GAAAAGG_10T/8A | ATTCT |
| 14.29 | ROUNDS_MDA5 | GAAAAAGG | 10T/8A | GAAAAAGG_10T/8A | ATTCT |
| 14.29 | ROUNDS_MDA5 | GAAAAGG | 9T/8A | GAAAAGG_9T/8A | ATTCT |
| 14.29 | ROUNDS_MDA5 | GAAAAGG | 9T/8A | GAAAAGG_9T/8A | ATTCT |
| 14.29 | ROUNDS_MDA5 | GAAAAGG | 9T/8A | GAAAAGG_9T/8A | ATTCT |
| 14.29 | ROUNDS_MDA5 | GAAAAGG | 10T/8A | GAAAAGG_10T/8A | ATTCT |
| 14.29 | ROUNDS_MDA5 | GAAAAGG | 10T/8A | GAAAAGG_10T/8A | ATTCT |
| 14.29 | ROUNDS_MDA5 | GAAAAGG | 10T/8A | GAAAAGG_10T/8A | ATTCT |
| 14.29 | ROUNDS_MDA5 | GAAAAAGG | 10T/7A | GAAAAAGG_10T/7A | ATTCT |
| 14.29 | ROUNDS_MDA5 | GAAAAAGG | 10T/8A | GAAAAAGG_10T/8A | ATTCT |
| 14.29 | ROUNDS_MDA5 | GAAAAGG | 10T/8A | GAAAAGG_10T/8A | ATTCT |
| 14.29 | ROUNDS_MDA5 | GAAAAAGG | 10T/8A | GAAAAAGG_10T/8A | ATTCT |
| 14.29 | ROUNDS_MDA5 | GAAAAGG | 10T/8A | GAAAAGG_10T/8A | ATTCT |

| | | | | | |
|---|---|---|---|---|---|
| 14.29 | ROUNDS_MDA5 | GAAAAGG | 10T/8A | GAAAAGG_10T/8A | ATTCT |
| 14.29 | ROUNDS_MDA5 | GAAAAGG | 10T/8A | GAAAAGG_10T/8A | ATTCT |
| 14.29 | ROUNDS_MDA5 | GAAAAGG | 10T/8A | GAAAAGG_10T/8A | ATTCT |
| 14.29 | ROUNDS_MDA5 | GAAAAGG | 10T/8A | GAAAAGG_10T/8A | ATTCT |
| 14.29 | ROUNDS_MDA5 | GAAAAGG | 10T/8A | GAAAAGG_10T/8A | ATTCT |
| 14.29 | ROUNDS_MDA5 | GAAAAAGG | 10T/7A | GAAAAAGG_10T/7A | ATTCT |
| 2.34 | ROUNDS_MDA5 | GAAAAGG | 10T/8A | GAAAAGG_10T/8A | ATTCT |
| 3.52 | ROUNDS_MDA5 | GAAAAGG | 10T/8A | GAAAAGG_10T/8A | ATTCT |
| 3.52 | ROUNDS_MDA5 | GAAAAGG | 10T/8A | GAAAAGG_10T/8A | ATTCT |
| 15.38 | ROUNDS_MDA5 | GAAAAGG | 10T/8A | GAAAAGG_10T/8A | ATTCT |
| 15.38 | ROUNDS_MDA5 | GAAAAGG | 10T/8A | GAAAAGG_10T/8A | ATTCT |
| 15.38 | ROUNDS_MDA5 | GAAAAGG | 10T/8A | GAAAAGG_10T/8A | ATTCT |
| 15.38 | ROUNDS_MDA5 | GAAAAGG | 10T/7A | GAAAAGG_10T/7A | ATTCT |
| 15.38 | ROUNDS_MDA5 | GAAAAGG | 10T/8A | GAAAAGG_10T/8A | ATTCT |
| 15.38 | ROUNDS_MDA5 | GAAAAAGG | 10T/8A | GAAAAAGG_10T/8A | ATTCT |
| 15.38 | ROUNDS_MDA5 | GAAAAGG | 10T/8A | GAAAAGG_10T/8A | ATTCT |
| 15.38 | ROUNDS_MDA5 | GAAAAAGG | 10T/8A | GAAAAAGG_10T/8A | ATTCT |
| 4.48 | ROUNDS_MDA5 | GAAAAAGG | 10T/8A | GAAAAAGG_10T/8A | ATTCT |
| 4.48 | ROUNDS_MDA5 | GAAAAGG | 10T/8A | GAAAAGG_10T/8A | ATTCT |
| 4.48 | ROUNDS_MDA5 | GAAAAAGG | 9T/7A | GAAAAAGG_9T/7A | ATTCT |
| 6.16 | ROUNDS_MDA5 | GAAAAAGG | 10T/7A | GAAAAAGG_10T/7A | ATTCT |
| 16.98 | ROUNDS_MDA5 | GAAAAGG | 10T/8A | GAAAAGG_10T/8A | ATTCT |
| 16.98 | ROUNDS_MDA5 | GAAAAGG | 10T/8A | GAAAAGG_10T/8A | ATTCT |
| 16.98 | ROUNDS_MDA5 | GAAAAGG | 10T/8A | GAAAAGG_10T/8A | ATTCT |
| 16.98 | ROUNDS_MDA5 | GAAAAAGG | 10T/8A | GAAAAAGG_10T/8A | ATTCT |
| 16.98 | ROUNDS_MDA5 | GAAAAGG | 10T/8A | GAAAAGG_10T/8A | ATTCT |
| 6.74 | ROUNDS_MDA5 | GAAAAGG | 10T/8A | GAAAAGG_10T/8A | ATTCT |
| 6.74 | ROUNDS_MDA5 | GAAAAGG | 10T/8A | GAAAAGG_10T/8A | ATTCT |
| 6.74 | ROUNDS_MDA5 | GAAAAAGG | 10T/8A | GAAAAAGG_10T/8A | ATTCT |
| 6.74 | ROUNDS_MDA5 | GAAAAGG | 10T/8A | GAAAAGG_10T/8A | ATTCT |
| 6.74 | ROUNDS_MDA5 | GAAAAGG | 10T/8A | GAAAAGG_10T/8A | ATTCT |
| 6.74 | ROUNDS_MDA5 | GAAAAGG | 10T/8A | GAAAAGG_10T/8A | ATTCT |
| 6.74 | ROUNDS_MDA5 | GAAAAGG | 10T/8A | GAAAAGG_10T/8A | ATTCT |
| 6.74 | ROUNDS_MDA5 | GAAAAGG | 10T/8A | GAAAAGG_10T/8A | ATTCT |
| 9.83 | ROUNDS_MDA5 | GAAAAAGG | 10T/8A | GAAAAAGG_10T/8A | ATTCT |
| 2.66 | ROUNDS_MDA8 | GAAAAGG | 10T/8A | GAAAAGG_10T/8A | ATTCT |
| 2.66 | ROUNDS_MDA8 | GAAAAGG | 10T/8A | GAAAAGG_10T/8A | ATTCT |
| 2.66 | ROUNDS_MDA8 | GAAAAGG | 10T/8A | GAAAAGG_10T/8A | ATTCT |
| 2.66 | ROUNDS_MDA8 | GAAAAAGG | 10T/8A | GAAAAAGG_10T/8A | ATTCT |
| 2.66 | ROUNDS_MDA8 | GAAAAGG | 10T/8A | GAAAAGG_10T/8A | ATTCT |
| 10.3 | ROUNDS_MDA8 | GAAAAAGG | 10T/7A | GAAAAAGG_10T/7A | ATTCT |
| 10.3 | ROUNDS_MDA8 | GAAAAGG | 10T/8A | GAAAAGG_10T/8A | ATTCT |
| 10.3 | ROUNDS_MDA8 | GAAAAGG | 10T/7A | GAAAAGG_10T/7A | ATTCT |
| 10.3 | ROUNDS_MDA8 | GAAAAAGG | 10T/8A | GAAAAAGG_10T/8A | ATTCT |
| 10.3 | ROUNDS_MDA8 | GAAAAGG | 10T/8A | GAAAAGG_10T/8A | ATTCT |
| 10.3 | ROUNDS_MDA8 | GAAAAAGG | 10T/8A | GAAAAAGG_10T/8A | ATTCT |

| | | | | | |
|---|---|---|---|---|---|
| 10.3 | ROUNDS_MDA8 | GAAAAGG | 10T/7A | GAAAAGG_10T/7A | ATTCT |
| 10.3 | ROUNDS_MDA8 | GAAAAGG | 10T/8A | GAAAAGG_10T/8A | ATTCT |
| 10.3 | ROUNDS_MDA8 | GAAAAGG | 10T/8A | GAAAAGG_10T/8A | ATTCT |
| 10.3 | ROUNDS_MDA8 | GAAAAAGG | 10T/8A | GAAAAAGG_10T/8A | ATTCT |
| 10.3 | ROUNDS_MDA8 | GAAAAGG | 10T/8A | GAAAAGG_10T/8A | ATTCT |
| 10.3 | ROUNDS_MDA8 | GAAAAGG | 10T/7A | GAAAAGG_10T/7A | ATTCT |
| 10.3 | ROUNDS_MDA8 | GAAAAAGG | 10T/7A | GAAAAAGG_10T/7A | ATTCT |
| 1.47 | ROUNDS_MDA8 | GAAAAGG | 10T/8A | GAAAAGG_10T/8A | ATTCT |
| 1.47 | ROUNDS_MDA8 | GAAAAGG | 10T/8A | GAAAAGG_10T/8A | ATTCT |
| 6.43 | ROUNDS_MDA8 | GAAAAGG | 10T/8A | GAAAAGG_10T/8A | ATTCT |
| 6.43 | ROUNDS_MDA8 | GAAAAGG | 10T/8A | GAAAAGG_10T/8A | ATTCT |
| 6.43 | ROUNDS_MDA8 | GAAAAGG | 10T/7A | GAAAAGG_10T/7A | ATTCT |
| 3.32 | ROUNDS_MDA8 | GAAAAAGG | 10T/7A | GAAAAAGG_10T/7A | ATTCT |
| 3.32 | ROUNDS_MDA8 | GAAAAGG | 10T/8A | GAAAAGG_10T/8A | ATTCT |
| 3.32 | ROUNDS_MDA8 | GAAAAGG | 10T/8A | GAAAAGG_10T/8A | ATTCT |
| 3.32 | ROUNDS_MDA8 | GAAAAGG | 10T/8A | GAAAAGG_10T/8A | ATTCT |
| 26.09 | ROUNDS_MDA8 | GAAAAGG | 10T/8A | GAAAAGG_10T/8A | ATTCT |
| 26.09 | ROUNDS_MDA8 | GAAAAAGG | 10T/7A | GAAAAAGG_10T/7A | ATTCT |
| 26.09 | ROUNDS_MDA8 | GAAAAGG | 10T/8A | GAAAAGG_10T/8A | ATTCT |
| 26.09 | ROUNDS_MDA8 | GAAAAAGG | 10T/8A | GAAAAAGG_10T/8A | ATTCT |
| 3.41 | ROUNDS_MDA8 | GAAAAGG | 10T/8A | GAAAAGG_10T/8A | ATTCT |
| 3.41 | ROUNDS_MDA8 | GAAAAGG | 10T/8A | GAAAAGG_10T/8A | ATTCT |
| 3.26 | ROUNDS_MDA8 | GAAAAGG | 10T/8A | GAAAAGG_10T/8A | ATTCT |
| 3.26 | ROUNDS_MDA8 | GAAAAGG | 10T/8A | GAAAAGG_10T/8A | ATTCT |
| 5.68 | ROUNDS_MDA8 | GAAAAGG | 10T/8A | GAAAAGG_10T/8A | ATTCT |
| 5.68 | ROUNDS_MDA8 | GAAAAAGG | 10T/7A | GAAAAAGG_10T/7A | ATTCT |
| 9.83 | ROUNDS_MDA5 | GAAAAGG | 10T/8A | GAAAAGG_10T/8A | ATTCT |
| 31.1 | ROUNDS_MDA5 | GAAAAGG | 10T/8A | GAAAAGG_10T/8A | ATTCT |
| 31.1 | ROUNDS_MDA5 | GAAAAGG | 10T/8A | GAAAAGG_10T/8A | ATTCT |
| 31.1 | ROUNDS_MDA5 | GAAAAGG | 10T/8A | GAAAAGG_10T/8A | ATTCT |
| 11.32 | ROUNDS_MDA8 | GAAAAAGG | 10T/7A | GAAAAAGG_10T/7A | ATTCT |
| 11.32 | ROUNDS_MDA8 | GAAAAGG | 10T/8A | GAAAAGG_10T/8A | ATTCT |
| 11.32 | ROUNDS_MDA8 | GAAAAGG | 10T/8A | GAAAAGG_10T/8A | ATTCT |
| 11.32 | ROUNDS_MDA8 | GAAAAAGG | 10T/8A | GAAAAAGG_10T/8A | ATTCT |
| 11.32 | ROUNDS_MDA8 | GAAAAAGG | 10T/7A | GAAAAAGG_10T/7A | ATTCT |
| 11.32 | ROUNDS_MDA8 | GAAAAGG | 10T/8A | GAAAAGG_10T/8A | ATTCT |
| 11.32 | ROUNDS_MDA8 | GAAAAGG | 10T/8A | GAAAAGG_10T/8A | ATTCT |
| 31.1 | ROUNDS_MDA5 | GAAAAGG | 10T/8A | GAAAAGG_10T/8A | ATTCT |
| 31.1 | ROUNDS_MDA5 | GAAAAGG | 10T/8A | GAAAAGG_10T/8A | ATTCT |
| 31.1 | ROUNDS_MDA5 | GAAAAGG | 10T/7A | GAAAAGG_10T/7A | ATTCT |
| 31.1 | ROUNDS_MDA5 | GAAAAAGG | 10T/8A | GAAAAAGG_10T/8A | ATTCT |
| 31.1 | ROUNDS_MDA5 | GAAAAAGG | 10T/8A | GAAAAAGG_10T/8A | ATTCT |
| 31.1 | ROUNDS_MDA5 | GAAAAGG | 10T/7A | GAAAAGG_10T/7A | ATTCT |
| 31.1 | ROUNDS_MDA5 | GAAAAGG | 10T/8A | GAAAAGG_10T/8A | ATTCT |
| 31.1 | ROUNDS_MDA5 | GAAAAGG | 10T/8A | GAAAAGG_10T/8A | ATTCT |
| 31.1 | ROUNDS_MDA5 | GAAAAGG | 10T/8A | GAAAAGG_10T/8A | ATTCT |

| | | | | | |
|---|---|---|---|---|---|
| 2.98 | ROUNDS_MDA8 | GAAAAGG | 10T/8A | GAAAAGG_10T/8A | ATTCT |
| 25 | ROUNDS_MDA8 | GAAAAGG | 10T/8A | GAAAAGG_10T/8A | ATTCT |
| 25 | ROUNDS_MDA8 | GAAAAGG | 10T/8A | GAAAAGG_10T/8A | ATTCT |
| 16.18 | ROUNDS_MDA8 | GAAAAAGG | 10T/8A | GAAAAAGG_10T/8A | ATTCT |
| 16.18 | ROUNDS_MDA8 | GAAAAAGG | 10T/8A | GAAAAAGG_10T/8A | ATTCT |
| 16.18 | ROUNDS_MDA8 | GAAAAGG | 10T/8A | GAAAAGG_10T/8A | ATTCT |
| 16.18 | ROUNDS_MDA8 | GAAAAAGG | 10T/8A | GAAAAAGG_10T/8A | ATTCT |
| 16.18 | ROUNDS_MDA8 | GAAAAGG | 10T/8A | GAAAAGG_10T/8A | ATTCT |
| 16.18 | ROUNDS_MDA8 | GAAAAAGG | 10T/8A | GAAAAAGG_10T/8A | ATTCT |
| 16.18 | ROUNDS_MDA8 | GAAAAGG | 10T/8A | GAAAAGG_10T/8A | ATTCT |
| 16.18 | ROUNDS_MDA8 | GAAAAGG | 10T/8A | GAAAAGG_10T/8A | ATTCT |

| CT1299 | RIGHT_FLANK | CT1299_flank | CT1291_FLAN | CT1291 | CT1291_Flank_comb. |
|---|---|---|---|---|---|
| 10C | | ATTCT_10C_ | AAATGGTCT | 10C | AAATGGTCT_10C |
| 10C | | ATTCT_10C_ | AAATGGTCT | 10C | AAATGGTCT_10C |
| 13C | | ATTCT_13C_ | AAATGGTCT | 9C | AAATGGTCT_9C |
| 10C | | ATTCT_10C_ | AAATGGTCT | 9C | AAATGGTCT_9C |
| 11C | | ATTCT_11C_ | AAATGGTCT | 9C | AAATGGTCT_9C |
| 10C | | ATTCT_10C_ | AAATGGTCT | 10C | AAATGGTCT_10C |
| 14C | | ATTCT_14C_ | AAATGGTCT | 9C | AAATGGTCT_9C |
| 10C | | ATTCT_10C_ | AAATGGTCT | 9C | AAATGGTCT_9C |
| 13C | | ATTCT_13C_ | AAATGGTCT | 10C | AAATGGTCT_10C |
| 13C | | ATTCT_13C_ | AAATGGTCT | 10C | AAATGGTCT_10C |
| 9C | | ATTCT_9C_ | AAATGGTCT | 8C | AAATGGTCT_8C |
| 13C | | ATTCT_13C_ | AAATGGTCT | 10C | AAATGGTCT_10C |
| 10C | | ATTCT_10C_ | AAATGGTCT | 9C | AAATGGTCT_9C |
| 11C | | ATTCT_11C_ | AAATGGTCT | 9C | AAATGGTCT_9C |
| 11C | | ATTCT_11C_ | AAATGGTCT | 9C | AAATGGTCT_9C |
| 14C | | ATTCT_14C_ | AAATGGTCT | 9C | AAATGGTCT_9C |
| 9C | | ATTCT_9C_ | AAATGGTCT | 8C | AAATGGTCT_8C |
| 10C | | ATTCT_10C_ | AAATGGTCT | 8C | AAATGGTCT_8C |
| 14C | | ATTCT_14C_ | AAATGGTCT | 8C | AAATGGTCT_8C |
| 10C | | ATTCT_10C_ | AAATGGTCT | 9C | AAATGGTCT_9C |
| 12C | | ATTCT_12C_ | AAATGGTCT | 9C | AAATGGTCT_9C |
| 10C | | ATTCT_10C_ | AAATGGTCT | 9C | AAATGGTCT_9C |
| 11C | | ATTCT_11C_ | AAATGGTCT | 9C | AAATGGTCT_9C |
| 10C | | ATTCT_10C_ | AAATGGTCT | 9C | AAATGGTCT_9C |
| 14C | | ATTCT_14C_ | AAATGGTCT | 8C | AAATGGTCT_8C |
| 11C | | ATTCT_11C_ | AAATGGTCT | 9C | AAATGGTCT_9C |
| 14C | | ATTCT_14C_ | AAATGGTCT | 10C | AAATGGTCT_10C |
| 11C | | ATTCT_11C_ | AAATGGTCT | 9C | AAATGGTCT_9C |
| 13C | | ATTCT_13C_ | AAATGGTCT | 11C | AAATGGTCT_11C |
| 11C | | ATTCT_11C_ | AAATGGTCT | 10C | AAATGGTCT_10C |
| 14C | | ATTCT_14C_ | AAATGGTCT | 10C | AAATGGTCT_10C |
| 14C | | ATTCT_14C_ | AAATGGTCT | 9C | AAATGGTCT_9C |
| 10C | | ATTCT_10C_ | AAATGGTCT | 9C | AAATGGTCT_9C |
| 10C | | ATTCT_10C_ | AAATGGTCT | 9C | AAATGGTCT_9C |
| 10C | | ATTCT_10C_ | AAATGGTCT | 9C | AAATGGTCT_9C |
| 11C | | ATTCT_11C_ | AAATGGTCT | 9C | AAATGGTCT_9C |
| 14C | | ATTCT_14C_ | AAATGGTCT | 10C | AAATGGTCT_10C |
| 10C | | ATTCT_10C_ | AAATGGTCT | 9C | AAATGGTCT_9C |
| 10C | | ATTCT_10C_ | AAATGGTCT | 9C | AAATGGTCT_9C |
| 9C | | ATTCT_9C_ | AAATGGT | 8C | AAATGGT_8C |
| 11C | | ATTCT_11C_ | AAATGGTCT | 10C | AAATGGTCT_10C |
| 11C | | ATTCT_11C_ | AAATGGTCT | 9C | AAATGGTCT_9C |
| 9C | | ATTCT_9C_ | AAATGGT | 8C | AAATGGT_8C |
| 9C | | ATTCT_9C_ | AAATGGT | 8C | AAATGGT_8C |
| 13C | | ATTCT_13C_ | AAATGGTCT | 9C | AAATGGTCT_9C |

| | | | | |
|---|---|---|---|---|
| 10C | ATTCT_10C_ | AAATGGTCT | 9C | AAATGGTCT_9C |
| 10C | ATTCT_10C_ | AAATGGTCT | 8C | AAATGGTCT_8C |
| 11C | ATTCT_11C_ | AAATGGTCT | 11C | AAATGGTCT_11C |
| 11C | ATTCT_11C_ | AAATGGTCT | 9C | AAATGGTCT_9C |
| 10C | ATTCT_10C_ | AAATGGTCT | 10C | AAATGGTCT_10C |
| 10C | ATTCT_10C_ | AAATGGTCT | 8C | AAATGGTCT_8C |
| 14C | ATTCT_14C_ | AAATGGTCT | 10C | AAATGGTCT_10C |
| 14C | ATTCT_14C_ | AAATGGTCT | 8C | AAATGGTCT_8C |
| 11C | ATTCT_11C_ | AAATGGTCT | 10C | AAATGGTCT_10C |
| 10C | ATTCT_10C_ | AAATGGTCT | 8C | AAATGGTCT_8C |
| 9C | ATTCT_9C_ | AAATGGTCT | 9C | AAATGGTCT_9C |
| 14C | ATTCT_14C_ | AAATGGTCT | 8C | AAATGGTCT_8C |
| 13C | ATTCT_13C_ | AAATGGTCT | 8C | AAATGGTCT_8C |
| 11C | ATTCT_11C_ | AAATGGTCT | 8C | AAATGGTCT_8C |
| 10C | ATTCT_10C_ | AAATGGTCT | 8C | AAATGGTCT_8C |
| 10C | ATTCT_10C_ | AAATGGTCT | 8C | AAATGGTCT_8C |
| 10C | ATTCT_10C_ | AAATGGTCT | 8C | AAATGGTCT_8C |
| 9C | ATTCT_9C_ | AAATGGT | 8C | AAATGGT_8C |
| 10C | ATTCT_10C_ | AAATGGT | 9C | AAATGGT_9C |
| 9C | ATTCT_9C_ | AAATGGT | 8C | AAATGGT_8C |
| 9C | ATTCT_9C_ | AAATGGT | 8C | AAATGGT_8C |
| 10C | ATTCT_10C_ | AAATGGT | 8C | AAATGGT_8C |
| 10C | ATTCT_10C_ | AAATGGTCT | 9C | AAATGGTCT_9C |
| 10C | ATTCT_10C_ | AAATGGTCT | 9C | AAATGGTCT_9C |
| 9C | ATTCT_9C_ | AAATGGT | 8C | AAATGGT_8C |
| 10C | ATTCT_10C_ | AAATGGT | 9C | AAATGGT_9C |
| 9C | ATTCT_9C_ | AAATGGTCT | 8C | AAATGGTCT_8C |
| 9C | ATTCT_9C_ | AAATGGT | 8C | AAATGGT_8C |
| 9C | ATTCT_9C_ | AAATGGT | 8C | AAATGGT_8C |
| 9C | ATTCT_9C_ | AAATGGT | 8C | AAATGGT_8C |
| 9C | ATTCT_9C_ | AAATGGT | 8C | AAATGGT_8C |
| 14C | ATTCT_14C_ | AAATGGTCT | 9C | AAATGGTCT_9C |
| 9C | ATTCT_9C_ | AAATGGT | 8C | AAATGGT_8C |
| 10C | ATTCT_10C_ | AAATGGTCT | 9C | AAATGGTCT_9C |
| 14C | ATTCT_14C_ | AAATGGTCT | 10C | AAATGGTCT_10C |
| 8C | ATTCT_8C_ | AAATGGTCT | 9C | AAATGGTCT_9C |
| 9C | ATTCT_9C_ | AAATGGTCT | 9C | AAATGGTCT_9C |
| 9C | ATTCT_9C_ | AAATGGTCT | 9C | AAATGGTCT_9C |
| 11C | ATTCT_11C_ | AAATGGTCT | 9C | AAATGGTCT_9C |
| 10C | ATTCT_10C_ | AAATGGTCT | 9C | AAATGGTCT_9C |
| 11C | ATTCT_11C_ | AAATGGTCT | 8C | AAATGGTCT_8C |
| 13C | ATTCT_13C_ | AAATGGTCT | 9C | AAATGGTCT_9C |
| 14C | ATTCT_14C_ | AAATGGTCT | 10C | AAATGGTCT_10C |
| 11C | ATTCT_11C_ | AAATGGTCT | 7C | AAATGGTCT_7C |
| 11C | ATTCT_11C_ | AAATGGTCT | 7C | AAATGGTCT_7C |
| 12C | ATTCT_12C_ | AAATGGTCT | 8C | AAATGGTCT_8C |

| | | | | | |
|---|---|---|---|---|---|
| 10C | | ATTCT_10C_ | AAATGGTCT | 9C | AAATGGTCT_9C |
| 12C | | ATTCT_12C_ | AAATGGTCT | 8C | AAATGGTCT_8C |
| 11C | | ATTCT_11C_ | AAATGGTCT | 7C | AAATGGTCT_7C |
| 10C | | ATTCT_10C_ | AAATGGTCT | 9C | AAATGGTCT_9C |
| 11C | | ATTCT_11C_ | AAATGGTCT | 7C | AAATGGTCT_7C |
| 12C | | ATTCT_12C_ | AAATGGTCT | 8C | AAATGGTCT_8C |
| 10C | | ATTCT_10C_ | AAATGGTCT | 9C | AAATGGTCT_9C |
| 10C | | ATTCT_10C_ | AAATGGTCT | 9C | AAATGGTCT_9C |
| 14C | | ATTCT_14C_ | AAATGGTCT | 11C | AAATGGTCT_11C |
| 11C | | ATTCT_11C_ | AAATGGTCT | 9C | AAATGGTCT_9C |
| 14C | | ATTCT_14C_ | AAATGGTCT | 11C | AAATGGTCT_11C |
| 11C | | ATTCT_11C_ | AAATGGTCT | 9C | AAATGGTCT_9C |
| 10C | | ATTCT_10C_ | AAATGGTCT | 8C | AAATGGTCT_8C |
| 14C | | ATTCT_14C_ | AAATGGTCT | 8C | AAATGGTCT_8C |
| 10C | | ATTCT_10C_ | AAATGGTCT | 9C | AAATGGTCT_9C |
| 10C | | ATTCT_10C_ | AAATGGTCT | 9C | AAATGGTCT_9C |
| 9C | | ATTCT_9C_ | AAATGGTCT | 9C | AAATGGTCT_9C |
| 10C | | ATTCT_10C_ | AAATGGTCT | 9C | AAATGGTCT_9C |
| 10C | | ATTCT_10C_ | AAATGGT | 9C | AAATGGT_9C |
| 12C | | ATTCT_12C_ | AAATGGTCT | 10C | AAATGGTCT_10C |
| 14C | | ATTCT_14C_ | AAATGGTCT | 10C | AAATGGTCT_10C |
| 10C | 5C-ATCAAA | ATTCT_10C_5 | AAATGGTCT | 9C | AAATGGTCT_9C |
| 11C | | ATTCT_11C_ | AAATGGTCT | 9C | AAATGGTCT_9C |
| 10C | | ATTCT_10C_ | AAATGGTCT | 10C | AAATGGTCT_10C |
| 10C | | ATTCT_10C_ | AAATGGTCT | 9C | AAATGGTCT_9C |
| 11C | | ATTCT_11C_ | AAATGGTCT | 9C | AAATGGTCT_9C |
| 11C | | ATTCT_11C_ | AAATGGTCT | 9C | AAATGGTCT_9C |
| 10C | | ATTCT_10C_ | AAATGGTCT | 9C | AAATGGTCT_9C |
| 14C | | ATTCT_14C_ | AAATGGTCT | 9C | AAATGGTCT_9C |
| 10C | | ATTCT_10C_ | AAATGGTCT | 10C | AAATGGTCT_10C |
| 14C | | ATTCT_14C_ | AAATGGTCT | 9C | AAATGGTCT_9C |
| 10C | | ATTCT_10C_ | AAATGGT | 8C | AAATGGT_8C |
| 13C | | ATTCT_13C_ | AAATGGTCT | 9C | AAATGGTCT_9C |
| 10C | | ATTCT_10C_ | AAATGGTCT | 10C | AAATGGTCT_10C |
| 10C | | ATTCT_10C_ | AAATGGTCT | 9C | AAATGGTCT_9C |
| 10C | | ATTCT_10C_ | AAATGGTCT | 9C | AAATGGTCT_9C |
| 10C | | ATTCT_10C_ | AAATGGTCT | 9C | AAATGGTCT_9C |
| 10C | | ATTCT_10C_ | AAATGGTCT | 9C | AAATGGTCT_9C |
| 10C | | ATTCT_10C_ | AAATGGTCT | 9C | AAATGGTCT_9C |
| 11C | | ATTCT_11C_ | AAATGGTCT | 10C | AAATGGTCT_10C |
| 13C | | ATTCT_13C_ | AAATGGTCT | 9C | AAATGGTCT_9C |
| 14C | | ATTCT_14C_ | AAATGGTCT | 10C | AAATGGTCT_10C |
| 13C | | ATTCT_13C_ | AAATGGTCT | 9C | AAATGGTCT_9C |
| 13C | | ATTCT_13C_ | AAATGGTCT | 10C | AAATGGTCT_10C |
| 10C | | ATTCT_10C_ | AAATGGTCT | 8C | AAATGGTCT_8C |
| 14C | | ATTCT_14C_ | AAATGGTCT | 9C | AAATGGTCT_9C |

| | | | | | |
|---|---|---|---|---|---|
| 10C | | ATTCT_10C_ | AAATGGTCT | 9C | AAATGGTCT_9C |
| 14C | | ATTCT_14C_ | AAATGGTCT | 10C | AAATGGTCT_10C |
| 11C | | ATTCT_11C_ | AAATGGTCT | 10C | AAATGGTCT_10C |
| 11C | | ATTCT_11C_ | AAATGGTCT | 10C | AAATGGTCT_10C |
| 12C | | ATTCT_12C_ | AAATGGTCT | 10C | AAATGGTCT_10C |
| 11C | | ATTCT_11C_ | AAATGGTCT | 11C | AAATGGTCT_11C |
| 14C | | ATTCT_14C_ | AAATGGTCT | 10C | AAATGGTCT_10C |
| 10C | | ATTCT_10C_ | AAATGGTCT | 9C | AAATGGTCT_9C |
| 14C | | ATTCT_14C_ | AAATGGTCT | 10C | AAATGGTCT_10C |
| 11C | | ATTCT_11C_ | AAATGGTCT | 9C | AAATGGTCT_9C |
| 11C | | ATTCT_11C_ | AAATGGTCT | 9C | AAATGGTCT_9C |
| 12C | | ATTCT_12C_ | AAATGGTCT | 9C | AAATGGTCT_9C |
| 10C | | ATTCT_10C_ | AAATGGTCT | 9C | AAATGGTCT_9C |
| 10C | | ATTCT_10C_ | AAATGGTCT | 9C | AAATGGTCT_9C |
| 10C | | ATTCT_10C_ | AAATGGTCT | 9C | AAATGGTCT_9C |
| 10C | | ATTCT_10C_ | AAATGGTCT | 9C | AAATGGTCT_9C |
| 12C | | ATTCT_12C_ | AAATGGTCT | 10C | AAATGGTCT_10C |
| 12C | | ATTCT_12C_ | AAATGGTCT | 10C | AAATGGTCT_10C |
| 12C | | ATTCT_12C_ | AAATGGTCT | 10C | AAATGGTCT_10C |
| 10C | | ATTCT_10C_ | AAATGGTCT | 9C | AAATGGTCT_9C |
| 10C | | ATTCT_10C_ | AAATGGTCT | 9C | AAATGGTCT_9C |
| 10C | | ATTCT_10C_ | AAATGGTCT | 9C | AAATGGTCT_9C |
| 14C | | ATTCT_14C_ | AAATGGTCT | 9C | AAATGGTCT_9C |
| 13C | | ATTCT_13C_ | AAATGGTCT | 10C | AAATGGTCT_10C |
| 14C | 5C-ATCAAA | ATTCT_14C_5 | AAATGGTCT | 9C | AAATGGTCT_9C |
| 14C | | ATTCT_14C_ | AAATGGTCT | 9C | AAATGGTCT_9C |
| 10C | | ATTCT_10C_ | AAATGGTCT | 9C | AAATGGTCT_9C |
| 13C | | ATTCT_13C_ | AAATGGTCT | 9C | AAATGGTCT_9C |
| 11C | | ATTCT_11C_ | AAATGGTCT | 9C | AAATGGTCT_9C |
| 13C | | ATTCT_13C_ | AAATGGTCT | 9C | AAATGGTCT_9C |
| 13C | | ATTCT_13C_ | AAATGGTCT | 9C | AAATGGTCT_9C |
| 14C | | ATTCT_14C_ | AAATGGTCT | 10C | AAATGGTCT_10C |
| 14C | | ATTCT_14C_ | AAATGGTCT | 10C | AAATGGTCT_10C |
| 14C | | ATTCT_14C_ | AAATGGTCT | 10C | AAATGGTCT_10C |
| 13C | | ATTCT_13C_ | AAATGGTCT | 9C | AAATGGTCT_9C |
| 14C | | ATTCT_14C_ | AAATGGTCT | 10C | AAATGGTCT_10C |
| 14C | | ATTCT_14C_ | AAATGGTCT | 9C | AAATGGTCT_9C |
| 14C | | ATTCT_14C_ | AAATGGTCT | 8C | AAATGGTCT_8C |
| 14C | | ATTCT_14C_ | AAATGGTCT | 9C | AAATGGTCT_9C |
| 14C | | ATTCT_14C_ | AAATGGTCT | 8C | AAATGGTCT_8C |
| 14C | | ATTCT_14C_ | AAATGGTCT | 8C | AAATGGTCT_8C |
| 14C | | ATTCT_14C_ | AAATGGTCT | 8C | AAATGGTCT_8C |
| 13C | | ATTCT_13C_ | AAATGGTCT | 11C | AAATGGTCT_11C |
| 10C | | ATTCT_10C_ | AAATGGTCT | 12C | AAATGGTCT_12C |
| 14C | | ATTCT_14C_ | AAATGGTCT | 11C | AAATGGTCT_11C |
| 14C | | ATTCT_14C_ | AAATGGTCT | 8C | AAATGGTCT_8C |

| | | | |
|---|---|---|---|
| 10C | ATTCT_10C_ | AAATGGTCT 9C | AAATGGTCT_9C |
| 11C | ATTCT_11C_ | AAATGGTCT 9C | AAATGGTCT_9C |
| 11C | ATTCT_11C_ | AAATGGTCT 9C | AAATGGTCT_9C |
| 10C | ATTCT_10C_ | AAATGGTCT 9C | AAATGGTCT_9C |
| 13C | ATTCT_13C_ | AAATGGTCT 9C | AAATGGTCT_9C |
| 11C | ATTCT_11C_ | AAATGGTCT 9C | AAATGGTCT_9C |
| 14C | ATTCT_14C_ | AAATGGTCT 9C | AAATGGTCT_9C |
| 12C | ATTCT_12C_ | AAATGGTCT 9C | AAATGGTCT_9C |
| 11C | ATTCT_11C_ | AAATGGTCT 9C | AAATGGTCT_9C |
| 13C | ATTCT_13C_ | AAATGGTCT 9C | AAATGGTCT_9C |
| 13C | ATTCT_13C_ | AAATGGTCT 9C | AAATGGTCT_9C |

| Serovar | pedersen_code |
|---|---|
| A | 3b.4.4 |
| A | 3b.4.4 |
| A | 3b.7.3 |
| A | 2b.4.3 |
| A | 3.5.3 |
| A | 3.4.4 |
| B | 3b.8.3 |
| Ba | 3b.4.3 |
| Ba | 3b.7.4 |
| Ba | 2.7.4 |
| A | 2b.3.2 |
| B | 3.7.4 |
| B | 3b.4.3 |
| B | 3b.5.3 |
| B | 3b.5.3 |
| Ba | 2.8.3 |
| A | 3b.3.2 |
| B | 3b.4.2 |
| B | 3b.8.2 |
| B | 3b.4.3 |
| A | 3b.6.3 |
| A | 3b.4.3 |
| B | 3.5.3 |
| B | 3.4.3 |
| Ba | 3b.8.2 |
| A | 3b.5.3 |
| A | 3b.8.4 |
| A | 3b.5.3 |
| A | 2b.7.5 |
| A | 3b.5.4 |
| A | 3b.8.4 |
| A | 2.8.3 |
| B | 3b.4.3 |
| B | 3b.4.3 |
| B | 3b.4.3 |
| Ba | 3b.5.3 |
| A | 3b.8.4 |
| Ba | 3b.4.3 |
| Ba | 2b.4.3 |
| B | 3b.3.2c |
| A | 3b.5.4 |
| B | 3b.5.3 |
| B | 3b.3.2c |
| Ba | 3.3.2c |
| A | 3b.7.3 |

| | |
|---|---|
| A | 3b.4.3 |
| A | 3b.4.2 |
| A | 3.5.5 |
| A | 3b.5.3 |
| A | 3.4.4 |
| A | 3b.4.2 |
| A | 2.8.4 |
| A | 3.8.2 |
| A | 3.5.4 |
| A | 3b.4.2 |
| A | 1a.3.3 |
| A | 3b.8.2 |
| A | 3b.7.2 |
| A | 1a.5.2 |
| A | 3b.4.2 |
| A | 3b.4.2 |
| B | 3.4.2 |
| B | 3b.3.2c |
| Ba | 3b.4.3c |
| B | 2.3.2c |
| Ba | 3b.3.2c |
| B | 3b.4.2c |
| B | 3b.4.3 |
| A | 3b.4.3 |
| B | 2.3.2c |
| B | 3.4.3c |
| B | 3.3.2 |
| Ba | 2.3.2c |
| B | 3b.3.2c |
| B | 3.3.2c |
| B | 3b.3.2c |
| B | 3b.8.3 |
| B | 3b.3.2c |
| B | 3b.4.3 |
| A | 3.8.4 |
| A | 1a.2.3 |
| A | 1a.3.3 |
| A | 1.3.3 |
| B | 3b.5.3 |
| Ba | 3b.4.3 |
| A | 3b.5.2 |
| A | 2.7.3 |
| A | 3.8.4 |
| A | 3b.5.8 |
| A | 3.5.8 |
| B | 3b.6.2 |

| | |
|---|---|
| Ba | 3b.4.3 |
| A | 3b.6.2 |
| A | 3b.5.8 |
| B | 3b.4.3 |
| A | 3b.5.8 |
| B | 2.6.2 |
| B | 3b.4.3 |
| B | 3b.4.3 |
| A | 3b.8.5 |
| A | 3b.5.3 |
| A | 3b.8.5 |
| A | 3b.5.3 |
| A | 2b.4.2 |
| A | 3b.8.2 |
| Ba | 3.4.3 |
| B | 3b.4.3 |
| B | 3.3.3 |
| Ba | 3.4.3 |
| B | 3b.4.3c |
| A | 1a.6.4 |
| A | 2.8.4 |
| Ba | 3b.4b.3 |
| A | 3b.5.3 |
| A | 3b.4.4 |
| B | 3.4.3 |
| A | 3b.5.3 |
| B | 3b.5.3 |
| A | 3b.4.3 |
| A | 3.8.3 |
| A | 3b.4.4 |
| A | 3b.8.3 |
| B | 3b.4.2c |
| A | 3b.7.3 |
| A | 3b.4.4 |
| B | 3.4.3 |
| A | 3b.4.3 |
| A | 3b.4.3 |
| A | 3b.4.3 |
| A | 3.4.3 |
| A | 3b.5.4 |
| A | 2.7.3 |
| A | 3b.8.4 |
| A | 2b.7.3 |
| A | 3.7.4 |
| Ba | 3b.4.2 |
| A | 3.8.3 |

| | |
|---|---|
| A | 2b.4.3 |
| A | 3b.8.4 |
| A | 3b.5.4 |
| A | 3.5.4 |
| A | 3b.6.4 |
| A | 2b.5.5 |
| A | 2.8.4 |
| A | 3b.4.3 |
| A | 3b.8.4 |
| Ba | 3b.5.3 |
| Ba | 3b.5.3 |
| A | 2b.6.3 |
| A | 2.4.3 |
| A | 3b.4.3 |
| A | 3b.4.3 |
| A | 3b.4.3 |
| A | 3b.6.4 |
| A | 2.6.4 |
| A | 3b.6.4 |
| A | 3.4.3 |
| A | 3b.4.3 |
| A | 3b.4.3 |
| A | 3b.8.3 |
| A | 3b.7.4 |
| Ba | 3b.8a.3 |
| A | 2.8.3 |
| B | 3b.4.3 |
| A | 3b.7.3 |
| A | 3b.5.3 |
| A | 3b.7.3 |
| A | 2.7.3 |
| A | 3b.8.4 |
| A | 3b.8.4 |
| A | 3.8.4 |
| A | 2.7.3 |
| A | 3b.8.4 |
| A | 3b.8.3 |
| A | 3b.8.2 |
| A | 3b.8.3 |
| A | 2b.8.2 |
| A | 3.8.2 |
| A | 3.8.2 |
| A | 2b.7.5 |
| A | 3b.4.7 |
| A | 3b.8.5 |
| A | 3b.8.2 |

| | |
|---|---|
| A | 3b.4.3 |
| Ba | 3b.5.3 |
| B | 3b.5.3 |
| B | 3.4.3 |
| A | 3.7.3 |
| B | 3b.5.3 |
| A | 3.8.3 |
| A | 3b.6.3 |
| B | 3.5.3 |
| A | 3b.7.3 |
| A | 3b.7.3 |
